# Metagenomic strain tracking reveals patterns of bacterial spread and the impact of water chlorination

**DOI:** 10.64898/2026.02.08.26345864

**Authors:** Daehyun D. Kim, Colin J. Worby, Hannah Wharton, Arjun Miklos, Benard Chieng, Sammy M. Njenga, Ashlee M. Earl, Amy J. Pickering

## Abstract

Bacterial infections are a major cause of morbidity and mortality among children under five in low- and middle-income countries (LMICs). Children in LMICs are exposed to and colonized by a range of pathogenic bacteria, yet patterns of bacterial exchange between humans are not well known, in part because culturing and sequencing single bacterial isolates is labor-intensive. Here, we apply a machine learning strain tracking approach to metagenomic data from 511 stool samples from children and mothers across urban and rural Kenyan communities to characterize bacterial dissemination and assess if community-wide water chlorination disrupts transmission. We identified distinct strain-sharing dynamics across species; potentially pathogenic taxa (e.g., *Escherichia*, *Enterococcus*, *Campylobacter*) exhibited distance-dependent dissemination driven by young children, while commensal taxa (e.g., *Bifidobacterium*, *Bacteroides*) showed patterns consistent with dietary exposure. Drinking water chlorination reduced community-level strain-sharing in rural communities. Our study provides the first strain-level insights into multi-species bacterial transmission dynamics in LMIC communities, identifying distinct dissemination pathways for facultative versus mostly anaerobic bacteria. Moreover, our findings highlight the utility of metagenomic strain tracking to uncover how community spread can be disrupted.

## Main

Bacterial enteric infections are a major cause of death globally. A study of 33 bacterial pathogens estimated they caused 13.6% of all global deaths and 56.2% of sepsis deaths annually, with the highest mortality rate in Sub-Saharan Africa.^1–3^ Children under five are particularly susceptible to diarrhea and growth faltering from bacterial infections; even asymptomatic carriage can impair nutrient absorption and immune development.^4^ Preventing child exposure is complicated by the presence of numerous reservoirs of bacterial pathogens across communities, including water, food, soil, domesticated livestock, and wildlife.^5^

To understand bacterial spatial dissemination patterns and the relative contributions of distinct transmission pathways, multi-species strain tracking is needed. However, strain tracking studies typically rely on culture-based methods that compare genome sequences from single bacterial isolates, offering only a narrow view of host microbial diversity and limited insight into microbial dynamics across populations.^6–8^ In contrast, metagenomic sequencing provides an untargeted, culture-independent view of microbial diversity;^9^ but attaining strain-level resolution is challenging due to the inherent complexity of microbiomes. Metagenomic strain tracking tools have recently been developed to enable culture-independent views of bacterial exchange between hosts and across adult social networks,^10–14^ but have yet to be used to assess how public health interventions could affect strain sharing.

Here, we use a machine-learning augmented metagenomic strain tracking approach^15^ developed by our team to study child strain sharing patterns in urban and rural Kenyan communities across a diverse range of bacterial organisms. We assess the spatial distance dependency of strain sharing and the relative contribution of different age groups in microbial dissemination, focusing on bacterial pathogens of public health concern (including *Escherichia coli*, *Campylobacter*, and *Enterococcus*) and contrasting observed patterns of dissemination with strictly anaerobic commensals. Further, we leverage a cluster-randomized control trial (cRCT) to quantify how community-scale water chlorination modulates bacterial strain sharing dynamics. Given drinking water is a major reservoir for bacterial pathogens in LMICs, our approach investigates the extent to which water treatment can perturb inter-personal strain sharing networks within and between households.^5,16,17^

## Results

### Overview of study designs

We analyzed 511 human stool samples collected from two studies in Kenya, one in Western Kenya and one in Nairobi. The Western Kenya study leveraged stool samples collected within a cRCT (the WASH Benefits Kenya trial) that evaluated the impact of a drinking water chlorination intervention on child health outcomes in rural communities (Fig. 1A and 1B).^18^ We analyzed 238 stool samples from 121 households across 10 clusters that received a chlorinated drinking water intervention and 10 geographically paired active control clusters (Fig. 1A). Stool samples were collected from each household two years post intervention delivery: one from a child enrolled in the study at birth (sampled at 19 – 29 months old; see materials and methods) and from an older child living in the same household (≥ 30 months old). In chlorinated water clusters, manual chlorine dispensers were installed at an average of five communal water points per cluster, and all compounds with children under five received bottled chlorine to treat collected rainwater.

**Fig. 1.**
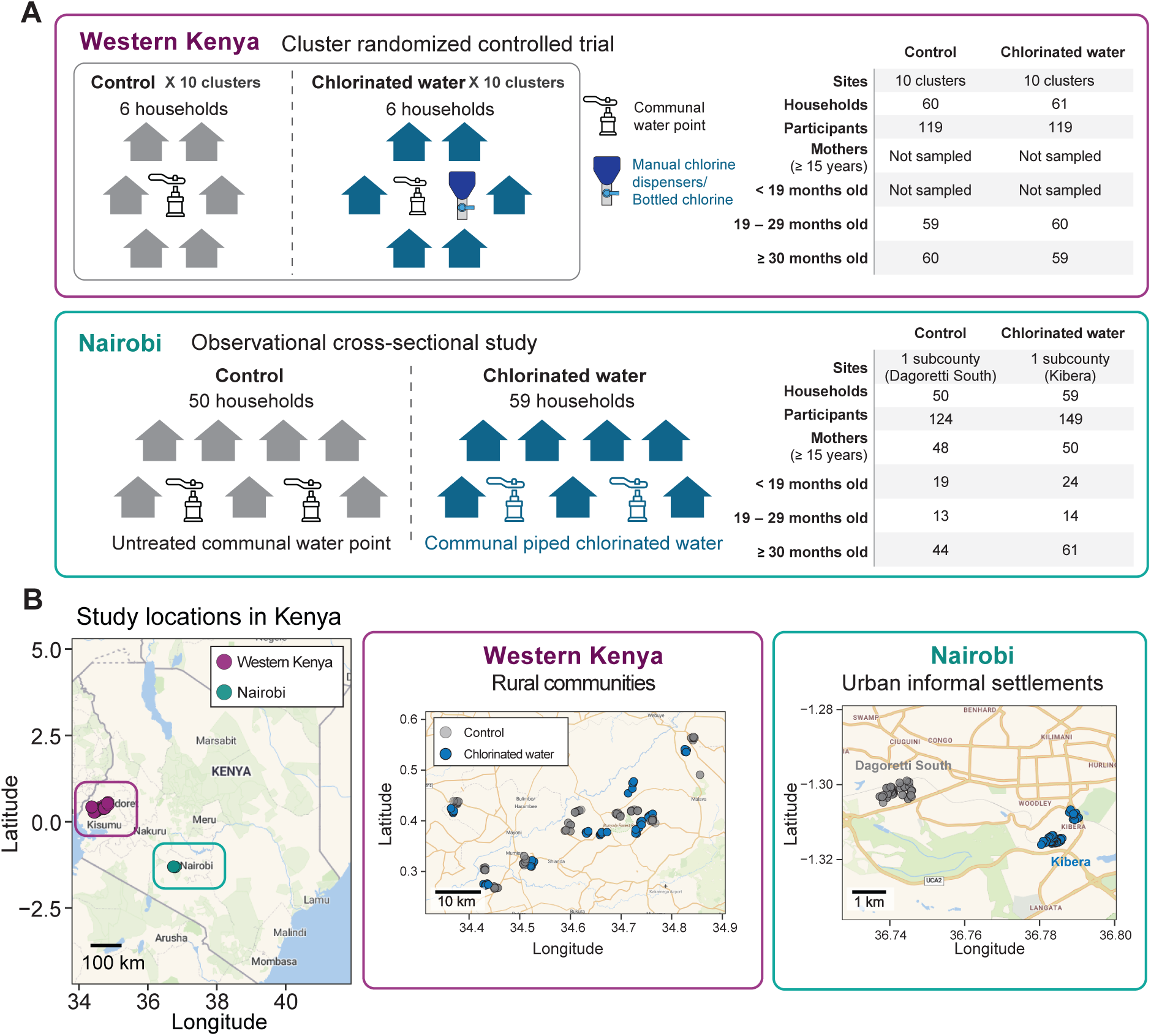
Overview of study design and locations in Kenya. (**A**) Schematic overview and summary of the Western Kenya and Nairobi study designs, including water treatment methods and participant characteristics. (**B**) Locations of the study sites and enrolled households in Western Kenya and Nairobi. Scale bars on the maps indicate the spatial distribution of households at different scales.

The Nairobi study was an observational cross-sectional study conducted in urban informal settlements across two counties: Dagoretti South (50 households) and Kibera (59 households) (Fig. 1A and 1B).^16^ Samples were collected from at least one child under the age of five in each household, with additional samples from siblings and mothers (≥ 15 years old) when applicable, totaling 273 samples from 109 households. Participants were categorized into four age groups: < 19 months old, 19 – 29 months old, ≥ 30 months old, and mothers (≥ 15 years old). Households in Kibera (chlorinated water group) collected their primary drinking water from communal taps supplied as piped chlorinated water, whereas Dagoretti South (control group) had no access to community-level chlorinated water. While not a randomized trial, the broader range of age groups included here provides greater insight into age- and relationship-dependent strain-sharing patterns.

### Characterization of gut microbial communities

We first conducted taxonomic profiling of the gut microbiota of all participants at the genus level.^19^ As expected, we observed significant clustering based on age group (Western Kenya: χ² = 66.6, *P* < 0.001; Nairobi: χ² = 215.3, *P* < 0.001; Fig. 2A), and increasing diversity with age (Fig. 2B), corresponding to the developmental phase of the gut microbiome.^20^ Compared to the Nairobi study, children in Western Kenya had lower gut diversity, but a higher *Prevotella:Bacteroides* ratio, potentially reflecting fiber-rich diets in rural communities (Fig. 2C and 2D).^21^

**Fig. 2.**
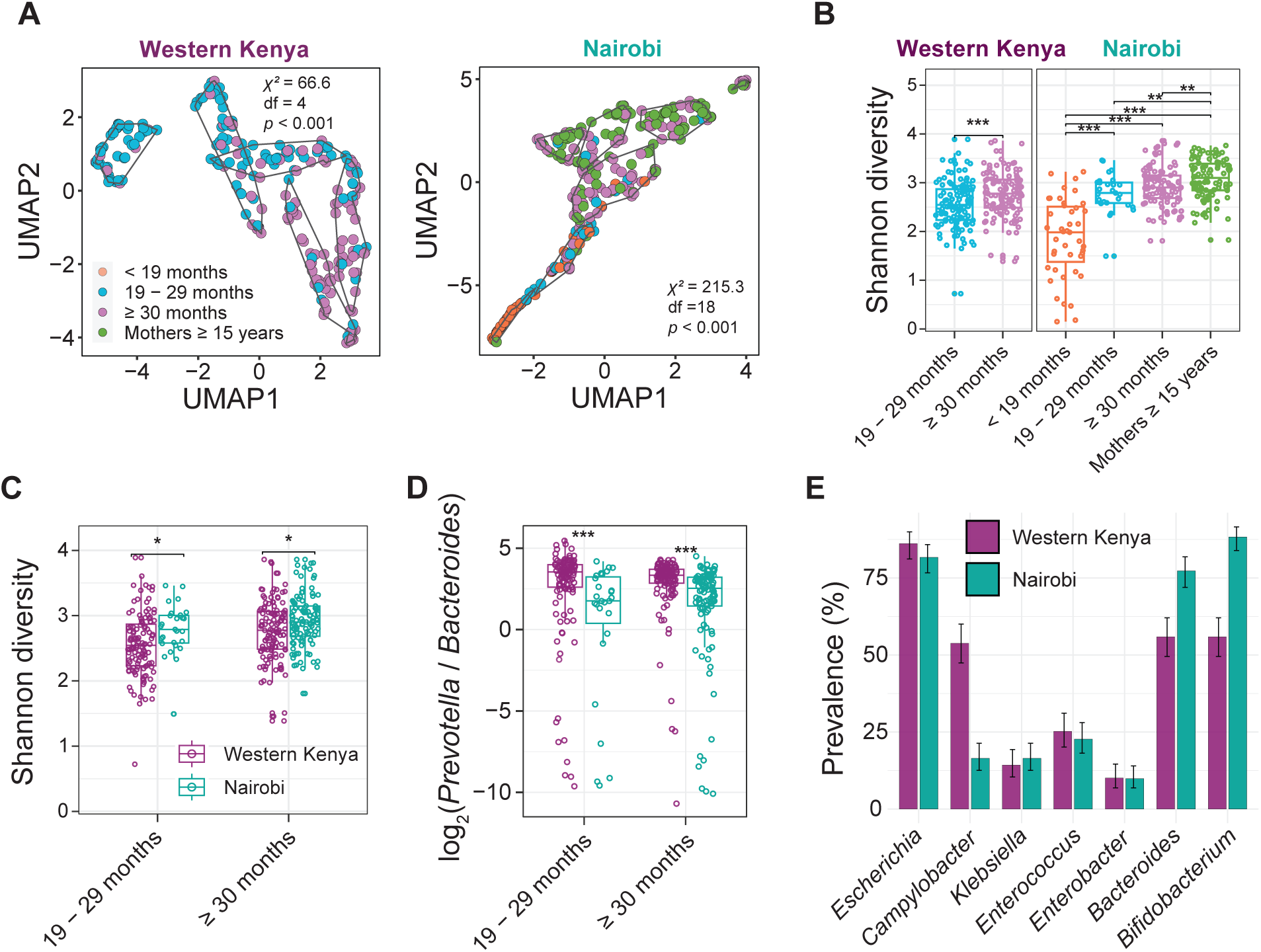
Characteristics of gut microbial communities. (**A**) Unsupervised clustering of microbial communities based on pairwise MinHash Jaccard distances. Differences in age group distributions across clusters were assessed using a two-sided chi-square test. (**B**) Shannon diversity of gut microbiota across age groups in Western Kenya and Nairobi. (**C**) Comparison of Shannon diversity between the two study sites, stratified by age group. (**D**) *Prevotella* to *Bacteroides* ratios between study sites, stratified by age group. Statistical comparisons were performed using two-sided Mann–Whitney U tests with Benjamini-Hochberg correction for multiple testing. (*: *P* < 0.05; **: *P* < 0.01; **: *P* < 0.001) (**E**) Prevalence of target bacterial species based on representative strain identification by StrainGE toolkit. Error bars indicate 95% confidence intervals calculated using the binomial proportion.

### Machine learning-driven strain-tracking enables detection of potential pathogenic and commensal strain sharing

We characterized potential pathogenic species present in each sample using StrainGE.^15^ We focused on genera of clinical interest, including the ESKAPEE pathogens (*Enterococcus*, *Staphylococcus*, *Klebsiella*, *Acinetobacter*, *Pseudomonas*, *Enterobacter*, and *Escherichia*), along with *Campylobacter* and *Salmonella*, which in this manuscript, we will collectively refer to as ‘pathogenic species’ (Fig. S1 and S2). Of these, *Escherichia* was the most prevalent in both studies; *Campylobacter* was notably more common in Western Kenya (53.8%, 128/238) than Nairobi (16.5%, 45/273) (Fig. 2E). *Pseudomonas*, *Salmonella*, *Acinetobacter,* and *Staphylococcus* strains were detected in at most one sample, consistent with their low relative abundances (< 0.57%; Fig. S3), which precluded them from being included in further analyses.

We calculated strain sharing rates of pathogenic species within- and between-households. Briefly, we first used StrainGE to assess strain genomic similarity between all pairs of samples with evidence of a strain match. Then using a machine learning classifier trained on datasets comprising longitudinal stool metagenomes, we identified sample pairs with instances of strain sharing – strain pairs with similarity concordant with that expected between strains collected from the same host (see materials and methods).

For comparison, we additionally evaluated strain sharing of abundant gut genera that, in contrast to pathogenic species, are largely anaerobic and typically commensal (herein referred to as ‘commensals’), and would be expected to exhibit distinct transmission dynamics (Fig. S4). We selected two high prevalence commensal genera, *Bifidobacterium* and *Bacteroides*, for strain-sharing analysis (Fig. 2E). Both are associated with vertical transmission from mother to child,^12,22–24^ which we hypothesized would result in strain sharing patterns distinct from pathogenic species – higher within-household sharing rates, and lower between-household sharing rates.

### Pathogenic species exhibit distance-dependent dissemination driven by young children

We identified a total of 322 instances of pathogenic species strain sharing in Western Kenya, and 155 in Nairobi. As expected, this was significantly elevated within households vs. between (*P* < 0.001, permutation test) (Fig. 3A and 3B). While the studies were not directly comparable due in part to the distinct age ranges of participants, we noted that pathogenic species strain sharing was higher in Western Kenya (Fig. 3B). Within households, this was primarily driven by high rates of *Campylobacter* strain sharing – among the 87 households in Western Kenya in which *Campylobacter* was detected, sharing was identified in 22 (25.3%), compared to 14.3% (5/35) of households in Nairobi (Fig. S5). Sharing of both *C. jejuni* and *C. infans* strains was identified; both species are associated with diarrhea in children and acquisition via animal feces.^25^ Between-household sharing of both *Campylobacter* and *Enterococcus* was also significantly higher in Western Kenya.

**Fig. 3.**
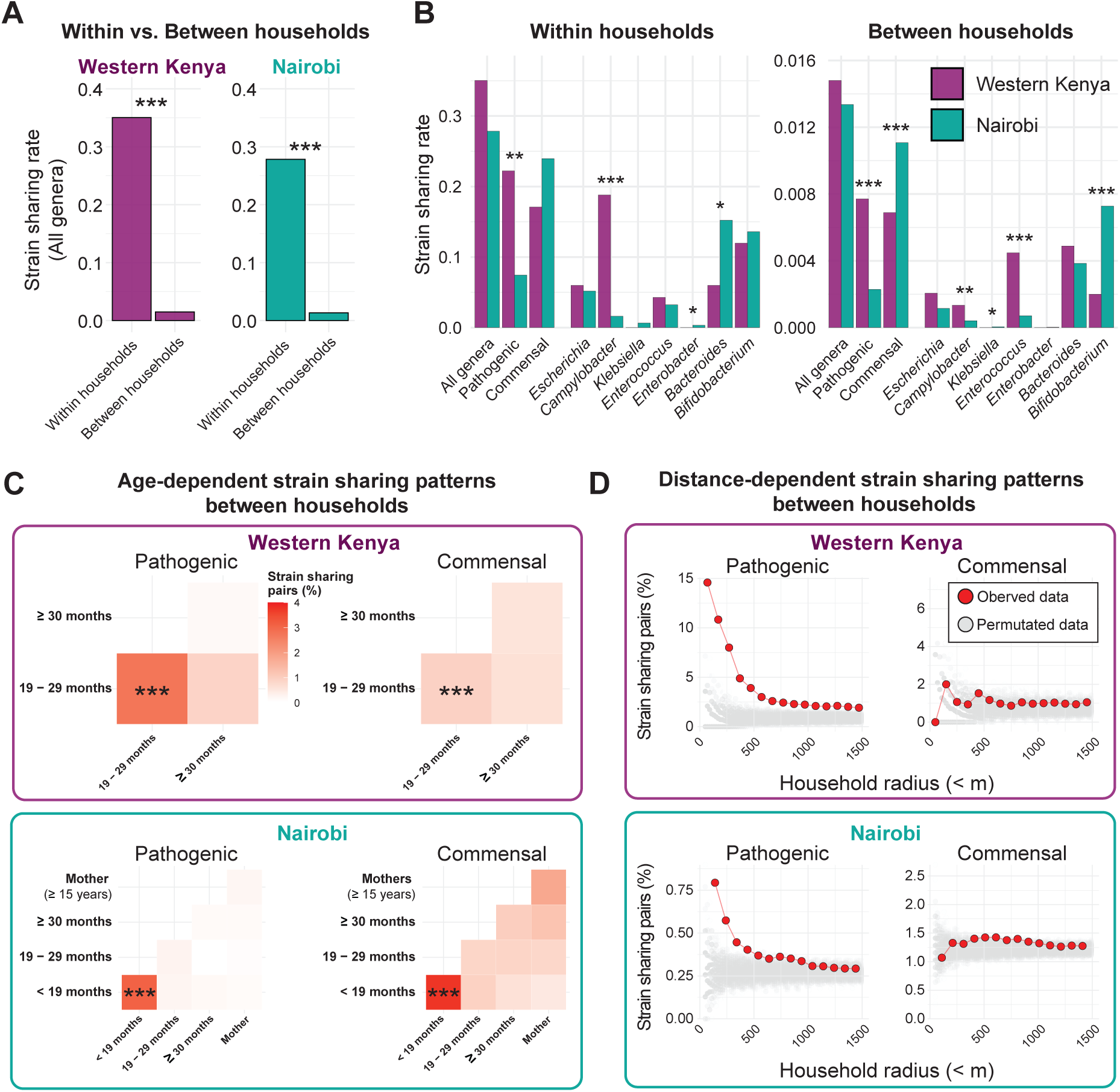
Bacterial strain-sharing patterns in two study sites. (**A**) Comparison of strain sharing rates within versus between households. Strain sharing rate was defined as the average proportion of total sample pairs (within- or between-household) that shared a strain. Statistical significance between groups was assessed using a two-sided non-parametric permutation test (1,000 iterations). (*: *P* < 0.05; **: *P* < 0.01; ***: *P* < 0.001). (**B**) Comparison of strain sharing rates between study sites. (**C**) Heatmaps of between-household strain sharing rates across age groups. Asterisks in tiles between the youngest age groups indicate significantly higher strain sharing rates compared to other age group pairs (***: *P* < 0.001). Full statistical test results are shown in Table S1. (**D**) Proportion of between-household strain sharing pairs across increasing household distance radius. Red points represent observed data; grey points show strain sharing proportions from 1,000 random permutations of household distances, representing the null expectation. Only household distances within 1,500m are shown.

To determine whether the elevated overall pathogenic species strain sharing rate observed in Western Kenya could be driven by differences in participant age, we considered only the 19 – 29 months and ≥ 30 months old age groups common to both studies. Strain sharing remained higher in Western Kenya, though the difference within households was not significant for this subset (Western Kenya: 0.22 cases per sample pair, Nairobi: 0.11; *P* = 0.085, permutation test) (Fig. S6). Pathogenic species were shared in the community most frequently among the youngest children. In Nairobi, pathogenic species were shared between households at significantly higher rates among children aged < 19 months (3.1% of pairs) compared to all other age groups (0.2% of pairs, *P* < 0.001, Fisher’s exact test) (Fig. 3C and 4A). The highest rates in this age group were observed for *Enterococcus*, specifically *E. avium* (Fig. 4B, S7), a species found across a range of animal populations.^26^ The Western Kenya study did not include this age group, though higher strain sharing rates were also observed in its youngest sampled age group (19 – 29 months; 1.8% vs. 0.4%, *P* < 0.001) (Fig. 3C and Fig. 4A). *E. hirae* was the most frequently shared species among these children; this organism is also found in animal populations where it can cause disease,^26^ as well as meat,^27^ and is only rarely associated with infections in humans (Fig. 4B, S8).^28^

**Fig. 4.**
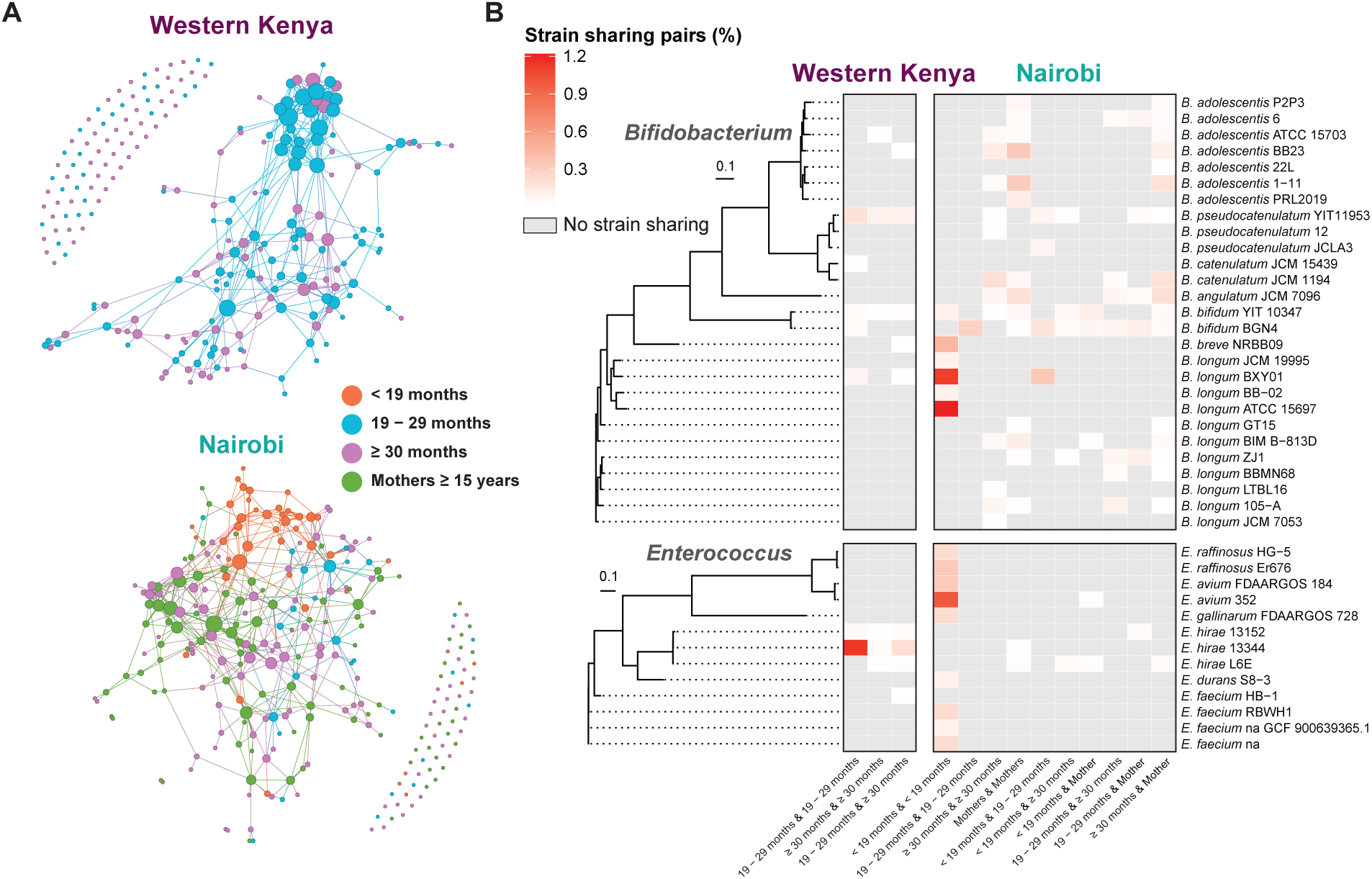
Between-household strain sharing patterns across age groups. (**A**) Individual-level between-household strain sharing networks in Western Kenya (left) and Nairobi (right). Each node represents an individual, colored by age group, with edges indicating detected strain sharing events. Node size is proportional to the number of strain sharing connections. Nodes with more connections are positioned closer together. (**B**) Heatmaps of representative *Bifidobacterium* and *Enterococcus* reference strains associated with between-household strain sharing. Phylogenetic trees were constructed using the maximum-likelihood method based on the alignment of core genes from the genomes of the representative reference strains. Between-household strain sharing rates for all target genera in each study are provided in Fig. S7 and S8.

To better understand between-household sharing, which could encompass a wide range of dynamics (e.g. transmission between neighbors, acquisition from shared local resources, or community-wide dissemination), we assessed the relationship between spatial relationships and strain sharing (Fig. 3D, S9). Strain sharing of pathogen species was significantly higher within clusters and villages vs. between (Western Kenya *P* = 0.009; Nairobi *P* = 0.027). Moreover, we found that pathogenic bacteria strain sharing rates decreased with distance between households; pathogenic strain sharing was significantly elevated below 500m in both settings, consistent with transmission occurring via close interpersonal contact or localized environmental contamination.^29^

### Commensal strain sharing in the community is consistent with dietary exposure

To contextualize our findings of pathogenic species strain sharing, we compared these patterns to commensal dynamics. As expected, these mostly anaerobic organisms were predominantly shared within households (*P* < 0.001, permutation test; Fig. 3B). Sampling of mothers in Nairobi allowed us to identify signals of vertical transmission, with *Bacteroides caccae* and *Bifidobacterium bifidum* in particular found to be frequently shared between mother and child (Fig. S10 and S11). Between-household commensal sharing was also identified, and similar to pathogenic species, this occurred most frequently among youngest children in both study sites; children aged < 19 months in Nairobi (3.8% of pairs vs. 1.0% of pairs across other age groups, *P* < 0.001) and children aged 19 – 29 months in Western Kenya (1.0% vs. 0.6%, *P* < 0.001) (Fig. 3C, 4A). In contrast to pathogenic species, commensals did not show distance-dependent strain sharing patterns between households (Fig. 3D), suggesting a common source of acquisition available across a larger area (e.g. food, milk, formula). Two *Bifidobacterium longum* reference genomes accounted for most strain sharing in the Nairobi study, observed almost exclusively among children aged < 19 months (Fig. 4B, S7). *B. longum* is a common colonizer of the infant gut, often acquired maternally,^11^ via food, or environmental contamination.^30,31^. However, we observed no sharing between mothers and their children of the two community *B. longum* strains. Finally, we assessed the possibility that observed between-household *Bifidobacterium* strain sharing could be driven by species or lineages with very low evolutionary rates, such that strains with distant common ancestors could still be classified as shared. In such a scenario, we would expect to see a comparable degree of strain sharing between the two study populations, which were close to independent (large geographic separation; no direct transmission, limited overlap in food distribution and resources); however, the between-study strain sharing rate was significantly lower than rates observed across each community (Fig. S12). Together, these results are consistent with dissemination in the community via dietary exposure.

### Water chlorination reduced strain sharing of pathogenic species in Western Kenya

To confirm that chlorinated water has minimal influence on healthy gut microbiome development in children,^32^ we examined overall gut microbial communities. Use of chlorinated water had limited impact on overall gut microbiome structure; there was no difference in diversity between chlorinated water and control groups (Fig. 5A), and unsupervised clustering based on metagenomic similarity revealed no association with chlorination in either study (Western Kenya: χ² = 1.8, *P* = 0.77; Nairobi: χ² = 10.5, *P* = 0.10) (Fig. 5B). A small number of taxa were differentially abundant between the groups, including *Megasphaera*, which was less abundant in the chlorinated water group in both studies, and *Klebsiella* and *Escherichia*, potentially pathogenic genera which exhibited increased abundance in the chlorinated water group (Fig. S13). These results are concordant with previous findings on the impact of water chlorination on children’s microbiomes.^32^

**Fig. 5.**
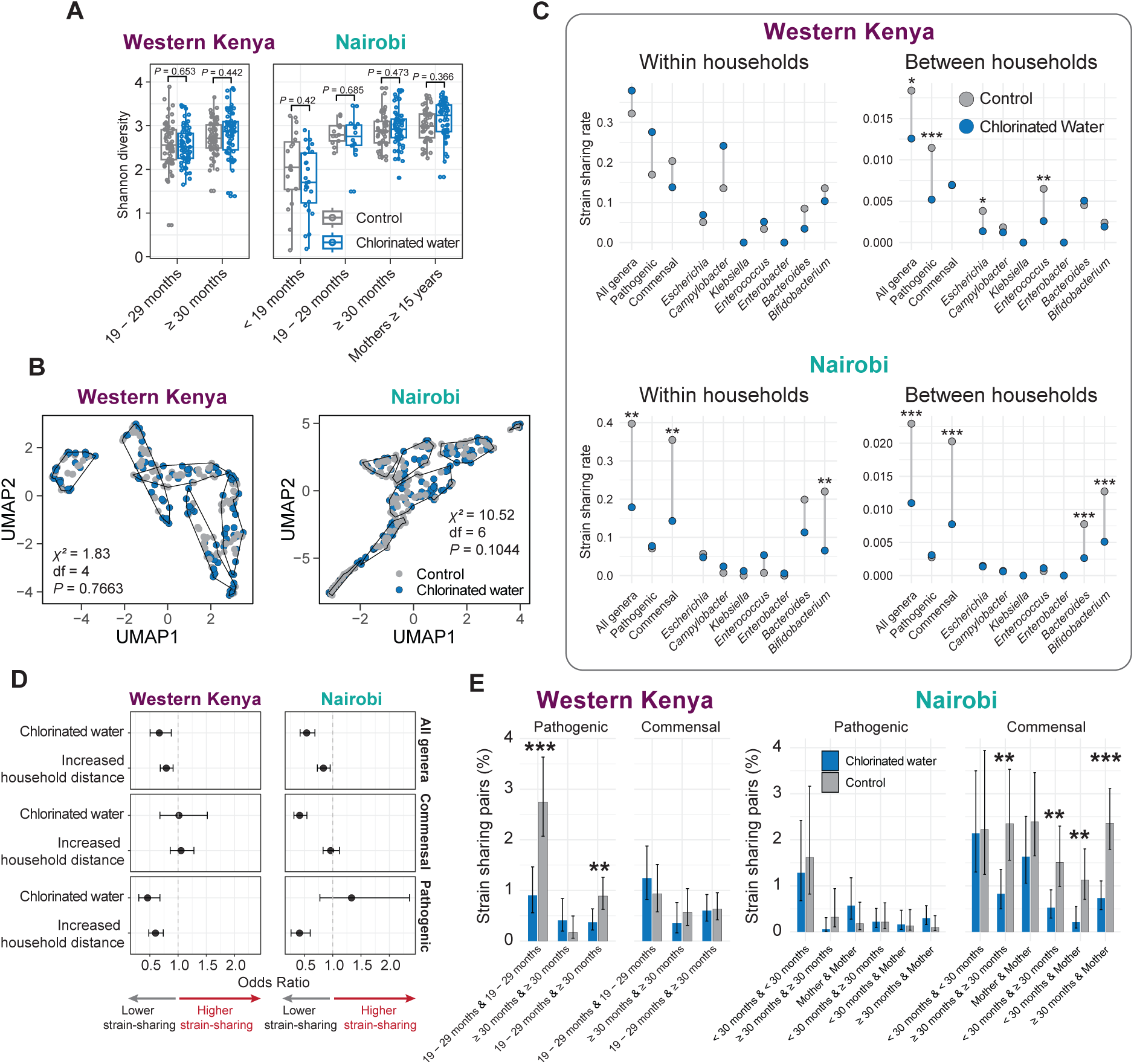
Effects of chlorinated water on overall gut microbiome and bacterial strain sharing patterns. (**A**) Shannon diversity of gut microbiomes stratified by age group. Statistical differences between control and chlorinated water groups were assessed using a two-sided Mann-Whitney U test. (**B**) Unsupervised clustering of microbial communities based on distance matrices calculated from pairwise MinHash Jaccard distances. Differences in sample distribution across clusters by treatment group were evaluated using a two-sided chi-square test. (**C**) Strain sharing rates within and between households by chlorinated water treatment. A two-sided non-parametric permutation test (1,000 iterations) was used to test statistical significance of differences in sharing rates between treatment groups. (*: *P* < 0.05; **: *P* < 0.01; ***: *P* < 0.001). (**D**) Odds ratios and 95% confidence intervals from logistic regression models evaluating the independent effects of chlorinated water and increased household distance on strain-sharing, adjusting for both variables. (**E**) Between-household strain sharing rates across age groups by access to chlorinated water. In Nairobi, age groups < 19 months and 19 – 29 months were combined into a single < 30 months group to improve statistical power. Error bars represent 95% confidence intervals based on binomial proportions. Statistical differences between treatment groups were assessed using two-sided Fisher’s exact tests (*: *P* < 0.05; **: *P* < 0.01; ***: *P* < 0.001).

We hypothesized that untreated water facilitates the spread of bacterial pathogens, and that community water chlorination reduces pathogen dissemination. We compared strain sharing based on water chlorination status to assess the impact of chlorinated water (Fig. 5C). Chlorinated water in both studies was associated with a significant reduction in overall strain sharing between households (Western Kenya *P* = 0.019; Nairobi *P* = 0.003, permutation test) (Fig. 5C); however, the impacted species differed. In Western Kenya, sharing of pathogenic species, including *Escherichia* (chlorinated water: 0.0014 cases per sample pair, control: 0.0038; *P* = 0.029) and *Enterococcus* (chlorinated water: 0.0026, control: 0.0065; *P* = 0.003), were significantly reduced. In contrast, in Nairobi, commensal strain sharing was reduced, driven by *Bifidobacterium* (chlorinated water: 0.0051, control: 0.013; *P* < 0.001) (Fig. 5C). Water chlorination did not impact pathogenic species sharing within households in either study; however, commensal strain sharing was reduced within households in Nairobi, driven again by *Bifidobacterium* (chlorinated water: 0.065, control: 0.22; *P* = 0.006) (Fig. 5C). Commensal dissemination was not impacted by water chlorination in Western Kenya.

To ensure the estimated effect of water chlorination was not confounded by the previously observed effects of household distance, we performed a logistic regression using water chlorination status and standardized household distance as predictors. Results were consistent with univariable observations; chlorinated water was associated with a reduced likelihood of strain sharing for pathogenic species in Western Kenya (OR = 0.46, 95% CI: 0.31–0.68, *P* < 0.001) and for commensal species in Nairobi (Odds Ratio [OR] = 0.41, 95% CI: 0.31–0.54, *P* < 0.001) (Fig. 5D).

Water chlorination significantly reduced pathogenic species strain sharing in Western Kenya in the youngest age group (chlorinated water: 0.9% of sample pairs; control: 2.7% of sample pairs; *P* < 0.001), though a similar effect was not seen in Nairobi (Fig. 5E). The reduction in strain sharing rates associated with chlorination treatment for commensal organisms appeared to be independent of age; strain sharing was lower in the water chlorination arm for all age group comparisons (Fig. 5E). Significant reductions were identified for age group comparisons with larger numbers of participants (mothers, children aged ≥ 30 months). A broad reduction of spatially-independent commensal strain sharing is concordant with decreased acquisition from community-wide sources, including food.

## Discussion

Our study provides some of the first insights into patterns of household and community bacterial strain sharing among children in LMIC settings. The metagenomic untargeted strain-tracking approach allowed us to uncover distinct patterns of bacterial dissemination across potentially pathogenic and commensal microorganisms. We observed pathogenic species strain sharing patterns to be consistent with highly localized, age-structured transmission networks;^12,13^ younger children were identified to be drivers of community-level spread. While this could reflect elevated levels of person-to-person transmission occurring within this age group, indirect transmission via common local reservoirs could also explain these patterns. For instance, infants and children have often been reported to play in feces-contaminated public spaces,^33,34^ ingesting soil.^35^ The frequent community-level sharing of *E. avium* and *E. hirae* observed in this study could be consistent with exposure to animal feces.^36,37^ *Enterococci* are hardy organisms and can survive desiccation in the environment,^38^ which could contribute to their elevated sharing rate. While *E. avium* and *E. hirae* are not a common cause of infection in humans, their burden in non-Western populations is not well characterized. Insights into transmission routes of these organisms highlight pathways that could also be utilized by higher risk pathogens such as *E. faecium* and *E. faecalis*.

While commensal strain sharing was enriched within-households, likely driven in part by vertical transmission,^23,39^ dissemination in the community had no spatial association, in contrast to pathogenic species. The relatively high rates of commensal strain sharing between households were unexpected, as these are largely anaerobes, thus having limited environmental transmissibility compared to oxygen-tolerant pathogenic species, which persist longer in the environment and spread more efficiently through pathways such as diarrhea-induced shedding.^12,40^ The lack of spatial association could reflect community-wide acquisition from common commercial food sources; *Bifidobacterium* is often found in fermented food or other dietary products,^30,31,41^ while *Bacteroides* has been detected in infant food in Kenya.^42^ Two *B. longum* strains, represented by reference genomes ATCC_15697 and BXY01,^43–45^ were responsible for most observed strain sharing in children aged < 19 months. Certain commensal strains, including the probiotic strain ATCC_15697 have also been reported to tolerate oxidative stress,^46–49^ potentially facilitating foodborne acquisition. The skew towards carriage and sharing among children aged < 19 months may reflect acquisition from common dietary sources consumed more frequently by this age group or maternal transmission of organisms originally acquired from food consumed by adults. Vertical transmission of these strains was not observed; however, this could have been missed due to undetectable levels of the strain in the mothers’ stool or carriage at alternative body sites – direct transmission could occur during vaginal delivery or breastfeeding.^50^ However, if this child group (aged < 19 months) did vertically acquire these commensal strains, the implied strain sharing between mothers across the community could still indicate a connection with dietary exposure.

In Western Kenya, the reduction in pathogenic species strain sharing in communities with water chlorination suggests disruption of local transmission networks. This could reduce the spread of clinically relevant pathogenic strains into households.^51,52^ Notably, the water chlorination intervention also reduced *E. coli* contamination in stored household drinking water^52^ and lowered child parasite infections^53^ in the parent WASH Benefits trial, but did not reduce self-reported diarrhea.^18^ The lack of an impact on diarrhea could be due to asymptomatic colonization, diverse etiologies and transmission pathways for diarrheal pathogens, or a need for complementary interventions.^54,55^ While no reduction of pathogenic species strain sharing was observed in Nairobi, this could be due to (i) a lower baseline level of pathogen strain sharing; (ii) differences in the relative importance of water as a driver of transmission between study settings;^56^ or (iii) missed confounders due to the observational study design in Nairobi. Nevertheless, our results give important insights on transmission dynamics for facultative versus strictly anaerobic bacteria.

Within-household pathogen strain sharing rates were not affected by chlorination in either study, suggesting within-household bacterial exchange is difficult to disrupt or overshadowing of water-mediated transmission pathways within households by alternative routes such as soil, hands, and food.^57^ Chlorinated water was associated with reduced commensal strain sharing in Nairobi. One potential mechanism for this could be the use of collected water in preparing foods, with chlorination inhibiting the proliferation of bacteria.^58^ Further research is needed to untangle commensal dynamics, and assess health impacts associated with reduced commensal sharing. Our study does have limitations. First, patterns of inferred strain sharing may not be fully reflective of transmission dynamics – it is possible that genomes may appear highly related despite hosts not sharing a recent common source. While there are likely some false positive strain sharing links identified, our benchmarking efforts suggested the false positive rate was low (see materials and methods). Our observed strain sharing patterns reflected expected dynamics; within-household rates were much higher than between household rates, pathogenic species sharing was distance dependent, and sharing rates between studies were observed to be extremely low, together affirming that strain sharing is capturing true patterns of microbial dissemination dynamics. Second, strain sharing events may be missed due to the limits of metagenomic detection. Notably, the microbiome becomes increasingly complex as children age,^59^ meaning that a given organism may achieve much lower coverage at the same abundance in a sample from an older *versus* younger host. This effect may contribute to the higher strain sharing rates observed between younger children in both studies; however, it would not affect conclusions related to the impact of chlorination or spatial dynamics. Finally, there were important differences between the two study sites, including in the implementation of water chlorination, study design, and participant demographics, which make comparisons between the two settings complex. Nevertheless, both studies independently showcase important, and previously uncharacterized bacterial dynamics, which will be a valuable foundation for future studies to quantify key transmission pathways and develop optimized interventions.

Through our metagenomic strain-tracking analysis, we uncover multi-species strain-level bacterial transmission dynamics in LMIC communities. We find pathogen spread is distance-dependent and driven by young children. Part of this may be mediated by exposure to shared spaces contaminated with animal fecal matter, future studies are needed to quantify transmission pathways from non-human hosts. Water chlorination reduced community-level pathogen sharing in the context of a randomized trial, providing important insights for the development and scale of optimized interventions to disrupt pathogenic bacterial exchange.

## Methods

### Study sites and sample collection

#### Western Kenya

We analyzed 238 child stool samples from 121 households across study sites in Kakamega, Bungoma, and Vihiga counties in rural western Kenya, collected as part of the WASH Benefits Kenya study.^18^ The WASH Benefits Kenya study was a cluster-randomized controlled trial that evaluated the impact of improved water, sanitation, handwashing, and nutrition on child diarrhea, linear growth, and early child development, with further details available in previous publications.^18^ In brief, geographic clusters of households with pregnant women in their second or third trimester were enrolled between November 2012 and May 2014; this birth cohort was followed until the study children reached approximately two years of age. Geographically adjacent clusters were block-randomized into one passive control, a double-sized active control group, and six intervention groups, including a water chlorination intervention. In water chlorination clusters, manual chlorine dispensers were installed at an average of five communal water points and bottled chlorine was delivered every 6 months to all compounds with children under five years old. Here, we leverage stool samples collected from children in the active control and water chlorination groups. After two years of intervention, stool samples were collected from the study child (at age 19 – 29 months) and an additional youngest older sibling (≥ 30 months) in the same compounds, with samples stored at −80 °C until use.

We randomly selected 20 clusters, consisting of 10 chlorinated drinking water intervention clusters geographically paired with 10 active control clusters, each containing at least six households. Within each cluster, we randomly chose six households and included stool samples from siblings in these households in our analysis.

#### Nairobi study

We collected 273 stool samples (175 from children and 98 from mothers) from 109 households located in urban informal settlements in Nairobi, Kenya, between June and August 2019.^60^ All enrolled households had at least one child under age 5 at the time of sampling and were located across two counties: Dagoretti South (50 households) and Kibera (59 households). Households in Kibera collected water from communal taps supplied by a chlorinated piped water system, as confirmed by chlorine residual testing at the tap during the study. In contrast, no communal taps in Dagoretti South were chlorinated.

We visited each household to collect information on demographics and household characteristics, including access to water, sanitation, and hygiene practices. Stool samples were collected from children under the age of five, with additional samples from older siblings (5 – 14 years) or mothers (≥ 15 years) in the same household if possible. These samples were further grouped into four age categories: infants (< 19 months), young children (19 – 29 months), older children (≥ 30 months), and mothers (15 ≥ years). Written informed consent for stool collection was obtained from each mother participant, along with child assent and parental consent for children. Approximately one week after the initial visit, we returned to the households to collect stool samples from household members. If stool samples were unavailable on the day of the visit, we made up to three additional attempts to collect them. All collected samples were stored at −80 °C until use.

All studies abided by relevant ethical regulations and guidelines. The original trial protocols for the Western Kenya study (WASH Benefits in Kenya) were approved by the committee for the protection of human subjects at the University of California, Berkeley (2011-09-3654), the institutional review board at Stanford University (IRB-23310), and the scientific and ethics review unit at the Kenya Medical Research Institute (SSC Protocol No. 2271). The Nairobi study received ethical approval from the Kenya Medical Research Institute (KEMRI) Scientific and Ethics Review Unit (KEMRI/SERU Protocol No. 3823) and the Tufts Health Sciences Institutional Review Board (13205). Additionally, a research permit was granted by the Kenyan National Commission for Science, Technology, and Innovation.

### DNA extraction and sequencing

Selected stool samples were retrieved from −80 °C and thawed on ice. DNA was then extracted from 0.2 g of each stool sample using the Qiagen QIAamp PowerFecal Pro DNA Kit (Qiagen, Hilden, Germany), following the manufacturer’s protocol. The concentration of the extracted DNA was measured using a Qubit 4.0 Fluorometer (Thermo Fisher Scientific, Waltham, MA), and the extracted DNA was shipped on dry ice in a cooler to the Broad Institute (Cambridge, MA).

Sequencing was performed on an Illumina NovaSeq 6000 generating 151 bp paired-end reads to yield a median of 11.8Gb per sample. Data were analyzed using the Broad Picard Pipeline (https://broadinstitute.github.io/picard). Reads were processed with KneadData to remove adapters and trim low base qualities, as well as to remove human-derived sequences.^61^

### Gut microbial community analysis

We analyzed the overall microbial compositions of stool samples to compare microbial communities across samples. The taxonomy of short reads in the sequence data was predicted using a combination of Kraken2 v2.1.2^62^ and Bracken v2.8,^19^ utilizing the Standard Kraken2/Bracken RefSeq indexes (5 June 2023 version) available from https://benlangmead.github.io/aws-indexes/k2. Reads unclassified by Kraken2 were excluded from the dataset, and read counts were aggregated at the genus level to avoid potential species-level misclassifications. In the aggregated genus-level abundance table, genera with a relative abundance of less than 0.01% were considered noise and filtered out.^63^ The resulting table was then used to calculate alpha diversity indices with the estimate_richness function from the phyloseq v1.42.0 package in R.^64^

We assessed microbiome similarities across samples using a k-mer-based sketch approach to incorporate large proportions of unclassified reads from Kraken2^62^ (approximately 50% on average). We used MASH v2.2.2^65^ to sketch k-mers from each sample’s sequence data and calculated pairwise MinHash Jaccard distances. The resulting distance matrix was dimensionally reduced using the uwot R package v0.2.2 (https://github.com/jlmelville/uwot), and samples were classified into clusters using unsupervised clustering with the Mclust R package v6.1.1.^66^ Genus-level differential abundance analysis was performed using the corncob R package v0.3.2, based on taxonomic profiles generated from Kraken2 results for genera with over 5% prevalence across samples and a minimum relative abundance of 0.1%.

### Bacterial strain identification

Our target bacterial strains in samples were identified using StrainGE v1.3.3,^15^ a k-mer-based strain-tracking tool designed to identify and compare bacterial strains within complex metagenomic data. Our targets included strains from genera of clinical importance, including ESKAPEE pathogens (*Enterococcus*, *Staphylococcus*, *Klebsiella*, *Acinetobacter*, *Pseudomonas*, *Enterobacter*, and *Escherichia*), as well as *Campylobacter* and *Salmonella*. For comparison, we also included *Bacteroides* and *Bifidobacterium*, which are largely anaerobes. To build the StrainGE database for each target genus, we downloaded all available complete genome sequences for each genus from the NCBI RefSeq database using the ‘ncbi-genome-download’ script. We constructed StrainGE databases for each genus using default settings, clustering reference genomes if they shared >99% of their k-mer content or had Jaccard similarity above 0.90. The number of genomes used in the database varied across genera, as follows: *Enterococcus* (337 representative genomes from 645 total reference genomes), *Staphylococcus* (382 from 1640), *Klebsiella* (552 from 2131), *Acinetobacter* (398 from 900), *Pseudomonas* (1058 from 1604), *Enterobacter* (323 from 564), *Escherichia* (863 from 2514), *Campylobacter* (321 from 573 total), *Salmonella* (183 from 1430), *Bacteroides* (70 from 80), and *Bifidobacterium* (142 from 225). Using StrainGST, the first module of StrainGE, we used quality-filtered short reads as inputs to identify the representative reference strains that were phylogenetically closest to the actual strains in each sample, along with their relative abundances.

We ran StrainGE on all stool metagenomes using each of the databases described above. After initially identifying representative StrainGE reference strains for each sample, we characterized genomic variants using bwa mem v0.7.17^67^ to align reads to the relevant references and the ‘straingr call’ command to filter variant positions. For sample pairs with common representative StrainGE references, strain comparison metrics were calculated using the ‘straingr compare’ function.

### Supplementing the StrainGE database for *Campylobacter*

Since the StrainGE databases comprised complete assembled genomes from cultured organisms, we sought to ensure there was sufficient representation for strains present in our samples. For each genus, we compared abundances from StrainGE with those from Kraken2; underestimation in StrainGE would likely arise from inadequate representation in its strain database. All genera were concordant with the exception of *Campylobacter*, with StrainGE predicting a lower abundance than Kraken2, suggesting the need to supplement the StrainGE database for *Campylobacter* (initially detected in 126 samples by StrainGE). To address this, we added two metagenome-assembled genomes (MAGs) of *Campylobacter* reconstructed from our samples to the StrainGE database (detected in 219 samples).

We assembled quality-controlled reads into contigs using MEGAHIT v1.2.9^68^ with default parameters. Contig depth coverage was calculated by mapping short reads onto the contigs using bwa mem v0.7.17^67^ with default parameters. The resulting contigs and depth data were then used as inputs for MetaBAT2 v2.15^69^ to reconstruct metagenome-assembled genomes (MAGs) for each sample. MAG quality was assessed with CheckM v1.2.1,^70^ retaining only those that met high-quality draft criteria (≥90% completeness and <5% contamination). The reconstructed MAGs across samples were dereplicated with dRep v3.4.5,^71^ using the ‘dereplicate’ command and parameters set as follows: ‘–S_algorithm fastANI –multiround_primary_clustering -ms 10000 -pa 0.9 -sa 0.95 -nc 0.30 -cm larger –completeness_weight 1 –contamination_weight 5 – strain_heterogeneity_weight 0 –N50_weight 0.5 –size_weight 1 –centrality_weight 200’. Taxonomy was assigned to the resulting set of MAGs with GTDB-tk v2.3.2^72^ ‘classify_wf’ using GTDB database release 214. Two high-quality *Campylobacter* MAGs meeting all requirements were retained, with contigs in each genome bin concatenated into a single sequence, bridged by 200 N’s to form a continuous sequence. These two genomes were then added to the existing genome set and processed using the same procedure to prepare the StrainGE database. After adding these MAGs, we found Kraken and StrainGE relative abundance estimates to be concordant.

### Identification of strain sharing across target genera

#### Characterizing within-host strain diversity

Previously, we used average callable nucleotide identity (ACNI), a measure of genomic similarity estimated by StrainGE, to determine whether *E. coli* strains present in different samples were plausibly ‘shared’ *i.e.*, similar enough to support a recent shared source of acquisition.^16^ We used a value of 99.95% similarity based on earlier benchmarking efforts.^15^ Since different species may be associated with varying levels of within-host diversity, the use of a universal genomic similarity cutoff to determine sharing is inappropriate. Thus, we sought to determine criteria for identifying strain sharing for a range of diverse gut organisms, most of which are considerably less well characterized than *E. coli*. To establish such criteria, we first attempted to characterize within-host strain diversity for each of the target genera. We assumed that strain diversity between hosts who share a common source of acquisition would be comparable to the strain diversity observed within a single host; as such, quantifying this metric from same-host metagenomic samples would guide threshold selection.^12^ Lacking repeated samples in this study, we utilized two external datasets comprising longitudinal stool metagenomes from (i) healthy international travelers (67 travelers with five samples over 1 year; Bioproject PRJNA528511),^73^ and (ii) cohorts of otherwise healthy women with and without recurrent urinary tract infections (UTIs) (31 participants with up to 21 samples over 1 year; Bioproject PRJNA31257).^74^

For each species, we selected a genomic similarity threshold which optimized sample classification as same-host or different-host based on maximization of the F1 score. This is analogous to a previous approach used to detect strain sharing across metagenomes using StrainPhlan,^12^ though here we opted to use StrainGE since this tool reports the presence of multiple strains within each species. We found ACNI thresholds varied widely between species (Fig. S14), likely due to a number of factors, including (i) differences in accumulated within-host diversity; (ii) differences in relative abundance; (iii) differences in exposures to new strains; (iv) differences in mutation rates; (v) the degree to which the strain database was representative of strains found in samples. Furthermore, it was not possible to determine robust ACNI thresholds for less prevalent (but often clinically relevant) species, limiting the utility of this approach in many settings.

#### Training a random forest model to identify strain-sharing

Given the between-species variation in ACNI threshold, and the relatively sparse data available for rarer species (particularly the pathogen target species), we developed a random forest model to discriminate between same-host and different-host sample pairs, with the ultimate goal of using this classifier to identify between-host strain sharing across multiple genera in our dataset. This approach removes the need to independently specify species similarity thresholds, while leveraging information across a range of organisms to classify sharing in rarer organisms, even those not included in the training set. Variables included in the model largely comprised output from StrainGE (Extended Data Table 1)

**Extended Data Table 1.** List of variables used to train a random forest model.

| Variable | Description |
| --- | --- |
| Genus | Genus of the target strain |
| Species | Species of the target strain |
| Length | Length of the genome |
| *Metrics generated by 'StrainGR compare' pairwise comparison* |  |
| commonPct | Percentage of genome positions callable in both samples |
| singlePct | Percentage of positions where both samples have a single strong call (i.e. no evidence for multiple alleles) |
| singleAgreePct<br>(ACNI) | Percentage of positions where both samples have single strong call, and the base call is the same. |
| sharedAllelesPct | Percentage of positions where both samples share an allele. This allows for positions to have multiple alleles, and at least one allele should match. |
| variantsPct | Percentage of positions where either sample has an allele other than the reference. |
| commonVariantPct | Percentage of variants where both samples share an allele |
| variantExactPct | Percentage of variants that are exactly the same in both samples (including the same positions with multiple alleles). |
| gapJaccardSimilarity | Jaccard similarity between samples of set of positions not marked as gap (i.e. analogous to gene content similarity). |

| *Sample metrics generated by ‘strainGR’ (averaged across sample pairs being compared)* |  |
| --- | --- |
| Mean coverage | Average depth of coverage along this genome. Includes multimapped reads, and multimapped reads are counted multiple times (for each alternative alignment location) |
| Mean callablePct | Percentage of positions in this genome with strong evidence for an allele (i.e. two good reads supporting a single allele) |
| Mean confirmedPct | Percentage of positions where there’s strong evidence for the reference allele (does not exclude positions with multiple alleles). |
| Mean snpPct | Percentage of positions with strong evidence for a single allele different from the reference. |
| Mean multiPct | Percentage of positions with strong evidence for multiple alleles (whether it includes the reference or not). |
| Mean lowmqPct | Percentage of positions where the majority of reads are mapped with low mapping quality, i.e. representing shared or repetitive regions. |
| Mean highPct | Percentage of positions with abnormally high coverage. |

We compared strain content in all pairs of samples in our external datasets, and aggregated StrainGE output for all sample pairs sharing a representative reference belonging to the target genus. We included the same target genera described above to train the model, and since many of the target pathogenic organisms were relatively rare in the training datasets, we included another, more abundant, facultative anaerobe, *Streptococcus*. In total, we identified representative StrainGE references belonging to 35 unique species within these genera in the training datasets.

We considered a total of 64,861 pairwise strain comparisons (both same-host and different-host) across samples in the traveler dataset, and 70,517 pairwise strain comparisons in the UTI dataset. We first trained random forest models on each dataset independently, using sklearn, by selecting a random 50% draw of pairwise strain comparisons from each dataset. We tested the performance of each random forest classifier to determine ‘same-host’ status on the remaining 50% of strain comparisons (‘test data’) from the corresponding dataset, as well as on the test data from the alternative dataset. Each classifier achieved an F1 score of 0.95-0.98 on its own dataset, with similar results when applying the classifier to the alternative dataset (F1=0.92).

We selected 50% of the 135,434 total pairwise strain comparisons across both external datasets to act as a training set, and used sklearn to establish a ‘combined’ random forest classifier. Applied to the remaining 50% of test data, this classifier achieved an F1 score of 0.96, considerably higher than the ACNI threshold approach (F1=0.78, Fig. S15). Performance was similarly improved for individual genera (Fig. S15). Finally, we used the classifier to determine strain sharing of ‘off-target’ species; those that were not included in the training data. We considered 13 species belonging to abundant genera which were not part of the training set (*Alistipes*, *Faecalibacterium*, *Blautia*), and sought to classify strain sharing across the 117,769 sample pairs with corresponding StrainGE hits. Across both datasets, the classifier achieved an F1 score of 0.89, again outperforming the ACNI cutoff approach on the same species (F1=0.70).

#### Applying the random forest classifier to Kenyan stool metagenomes

Finally, we applied the combined random forest classifier to the aggregated pairwise strain comparison metrics from the Kenyan stool metagenomes. Machine learning predictions showed clear separation in estimated nucleotide identity between shared and non-shared strains (Fig. S16). Average strain-sharing rates were defined as the proportion of total sample pairs (all within-household pairs or all between-household pairs) exhibiting strain-sharing.

We built strain-sharing networks and calculated network statistics, using the igraph v2.0.2 package^75^ in R. The networks were visualized using Gephi v0.10^76^ with the Force Atlas layout.

### Calculation of strain-sharing rate by household distance radius

To investigate how household distances affect strain-sharing, we calculated distances between households using GPS coordinates with the geosphere v1.5^77^ package in R. Across a household distance range of 50 m to 1500 m, in increments of 10 m, we calculated the percentage of sample pairs between households sharing any bacterial strain within each distance threshold. To assess whether the observed pattern between strain-sharing and household distance was non-random, we compared the actual data with 1,000 permuted strain-sharing percentage profiles, recalculated by randomly permuting household distances among all sample pairs.

### Statistical analysis

All statistical analyses were conducted in R version 4.2.1.72. Data normality was assessed before applying relevant statistical tests. Alpha diversity of microbial communities was compared between groups using pairwise Mann-Whitney U tests. The distribution of samples across microbial community composition clusters was assessed using a chi-squared test. For strain sharing analysis, we calculated household-level strain sharing rates as the average number of strain sharing sample pairs, normalized by the total number of sample pairs within each household or between household pairs. We assessed differences in mean household-level strain sharing rates using a two-sided nonparametric permutation test with 1,000 resampling iterations. To compare the percentage of individual-level strain sharing pairs between groups, we used a two-sided Fisher’s exact test. Additionally, to compare the effects of water chlorination and household distance on strain sharing, we applied a logistic regression model with the glm function in R to evaluate the significance and magnitude of the coefficients for these two variables.

## Data availability

All raw sequence data were deposited in the NCBI database under BioProject accession number PRJNA1356541. The external longitudinal stool metagenomic datasets utilized for a random forest model training are publicly available in the NCBI Sequence Read Archive under BioProject PRJNA528511 and BioProject PRJNA31257.

## Code availability

R scripts and Linux shell scripts used for the analysis are available at https://github.com/danielkim617/bacterial-strain-sharing-patterns-and-the-impact-of-water-chlorination

## Acknowledgments

This work was financially supported by National Institutes of Health grants 5R21AI171890 (AJP, AME) and U19AI110818 (AME), National Science Foundation grant 2143622 (AJP), and Bill and Melinda Gates Foundation grant OPP1200651and OPPGD759 (AJP, SMN). AJP is a Biohub, San Francisco Investigator. We thank Clair Null, John Colford, Christine Stewart, Benjamin Arnold, Maya Nadimpalli, Jenna Swarthout, and John Mboya for their role in the studies that collected the samples analyzed in this manuscript.

## Author contributions

AJP, AME, DDK, CJW, and SMN conceptualized the study. Methodology was developed by DDK, CJW, HW, and AM. Investigation was conducted by DDK and AJP. Data visualization was carried out by DDK and CJW. Funding acquisition was secured by AJP, AME, and SMN. DDK, CJW, AJP, and AME wrote the first draft of the manuscript. All authors edited the final manuscript. AJP, AME, and SMN supervised the project.

## Competing interests

The authors declare no competing interests.

## Notes

### Competing Interest Statement

The authors have declared no competing interest.

### Clinical Trial

NCT01704105

### Author Declarations

The original trial protocols for the Western Kenya study (WASH Benefits in Kenya) were approved by the committee for the protection of human subjects at the University of California, Berkeley (2011-09-3654), the institutional review board at Stanford University (IRB-23310), and the scientific and ethics review unit at the Kenya Medical Research Institute (SSC Protocol No. 2271). The Nairobi study received ethical approval from the Kenya Medical Research Institute (KEMRI) Scientific and Ethics Review Unit (KEMRI/SERU Protocol No. 3823) and the Tufts Health Sciences Institutional Review Board (13205).

