## Supplemental Information for "Metagenomic strain tracking reveals patterns of bacterial spread and the impact of water chlorination"

**Table of Contents:**

**Supplementary Note**
**Supplementary Figures (Fig. S1–S16)**

### Supplementary Note

### Reporting on sex

In the Western Kenya study, 43.9% of children (50 of 119) in control clusters were female, and 41.4% (48 of 119) in clusters with access to chlorinated water were female. In the Nairobi study, 43.4% of child participants (33 of 76) in control clusters and 57.6% (57 of 99) in clusters with access to chlorinated water were female. All participating mothers in the Nairobi study were female.

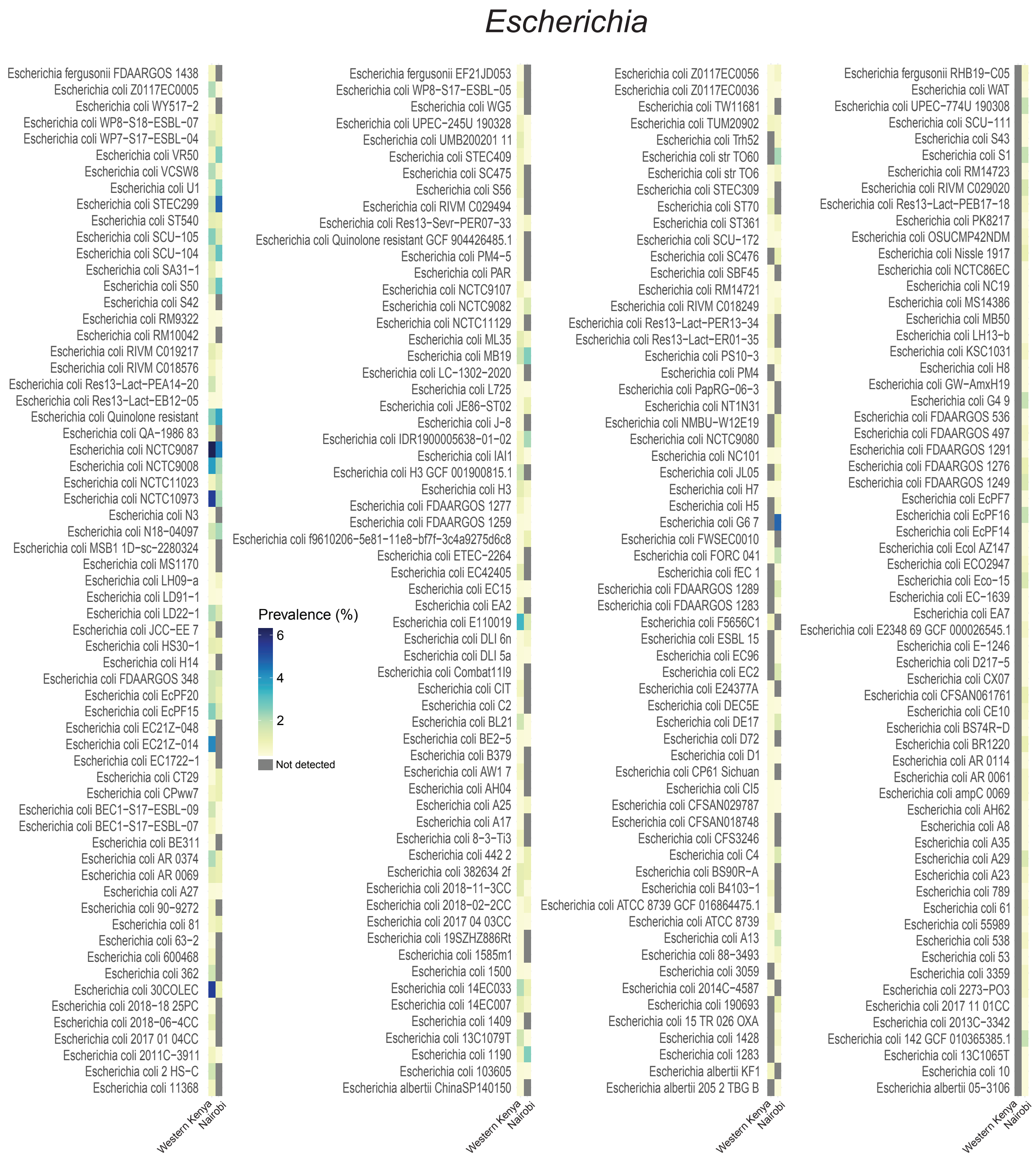


**Fig. S1. StrainGE-identified representative *Escherichia* reference strains.** Prevalence was calculated as the proportion of samples in which each strain was detected out of the total number of samples in each study site.

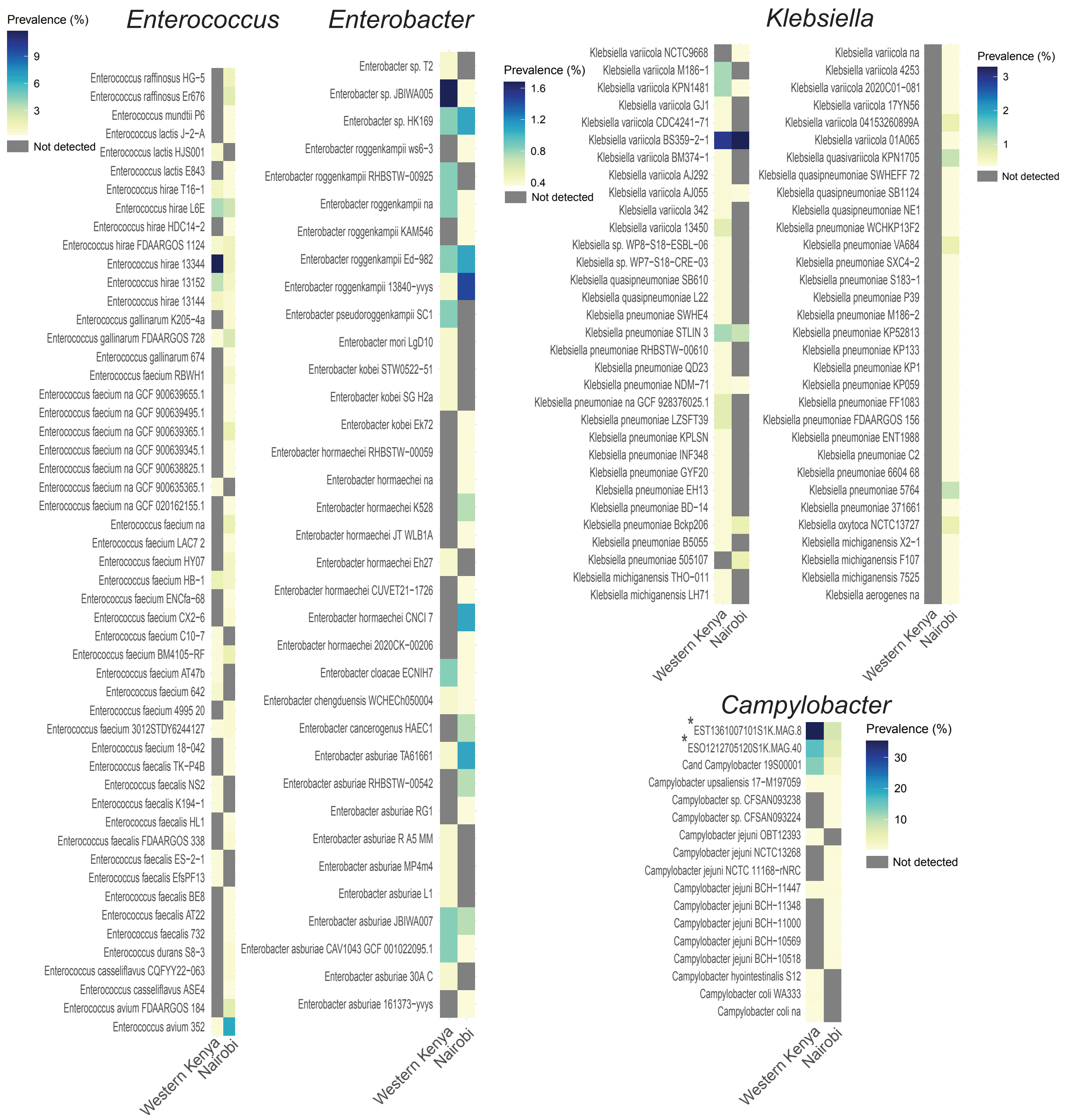


**Fig. S2. StrainGE-identified representative reference strains of *Klebsiella*, *Enterococcus*, *Enterobacter*, and *Campylobacter*.** Prevalence was calculated as the proportion of samples in which each strain was detected out of the total number of samples in each study site. *Campylobacter* strains marked with an asterisk indicate metagenome-assembled genomes reconstructed in this study.

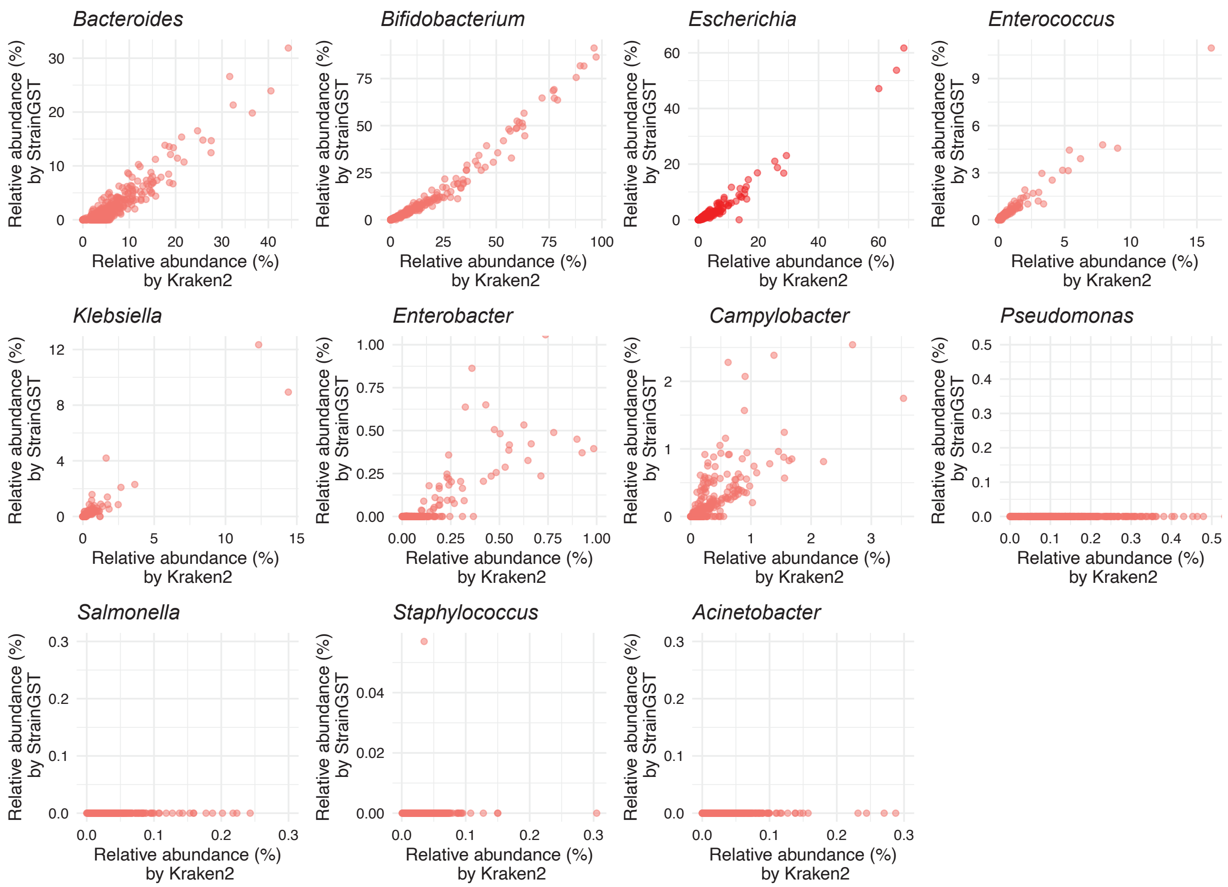


**Fig. S3. Relative abundances of target bacterial genera estimated using StrainGE and Kraken2.**

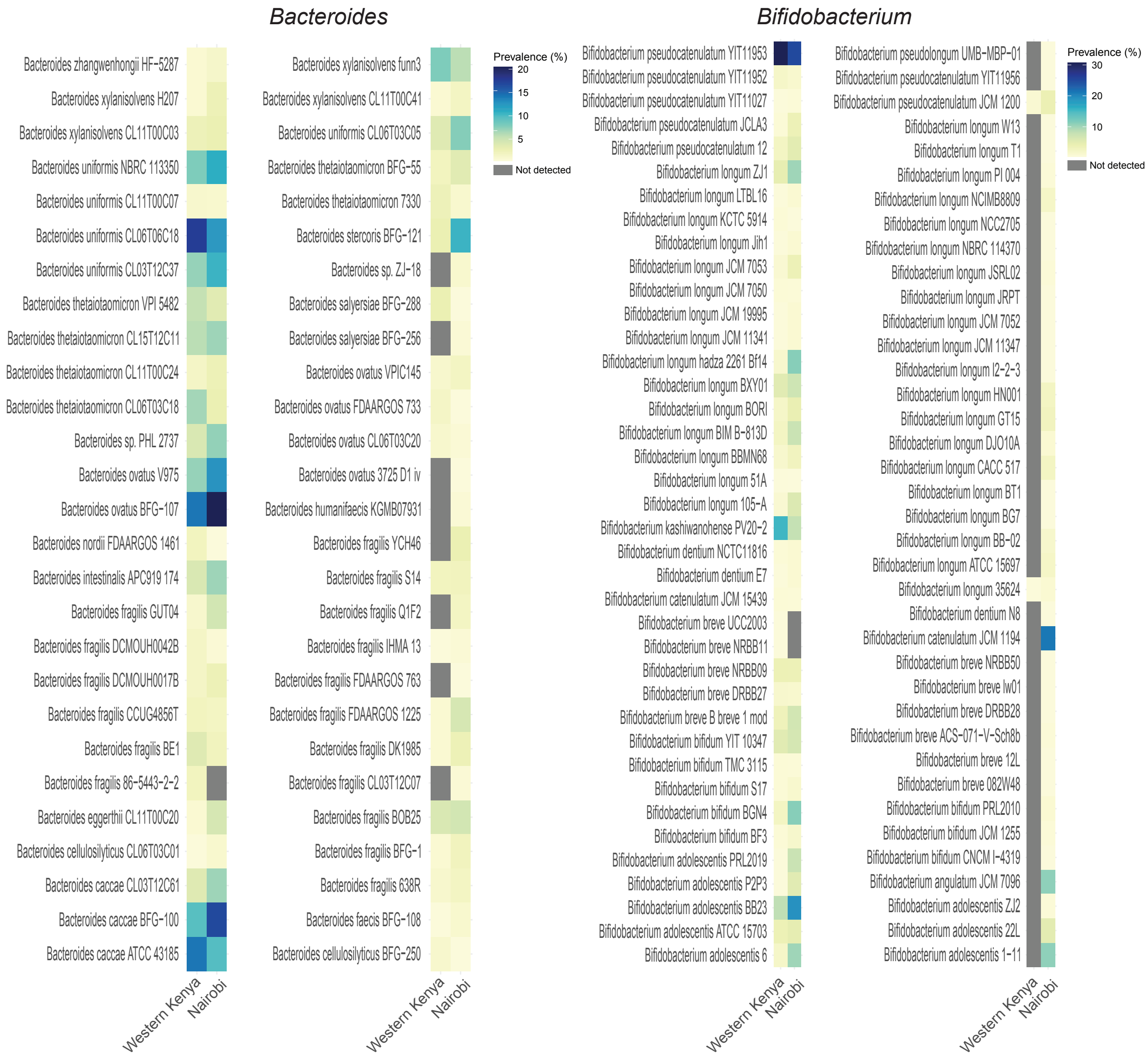


**Fig. S4. StrainGE-identified representative reference strains of *Bacteroides* and *Bifidobacterium*.** Prevalence was calculated as the proportion of samples in which each strain was detected out of the total number of samples in each study site.

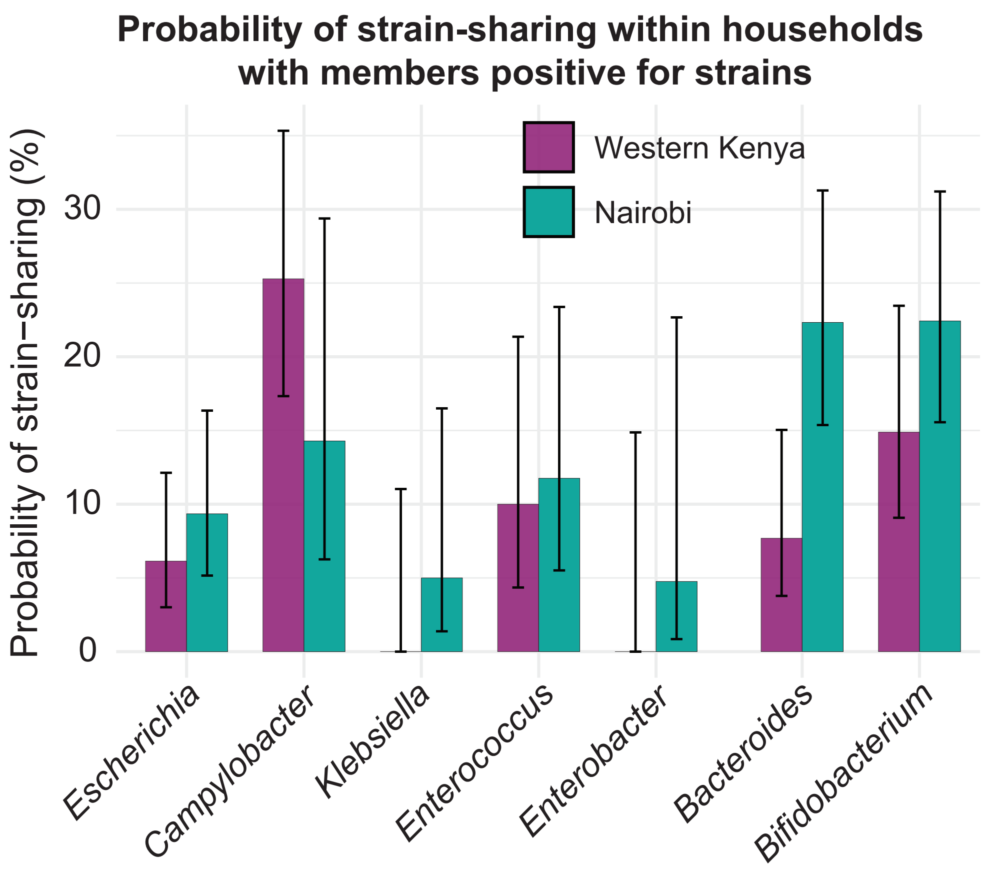


**Fig. S5. Probability of strain-sharing within households with at least one member positive for strains.** Error bars indicate 95% confidence intervals calculated using the binomial proportion.

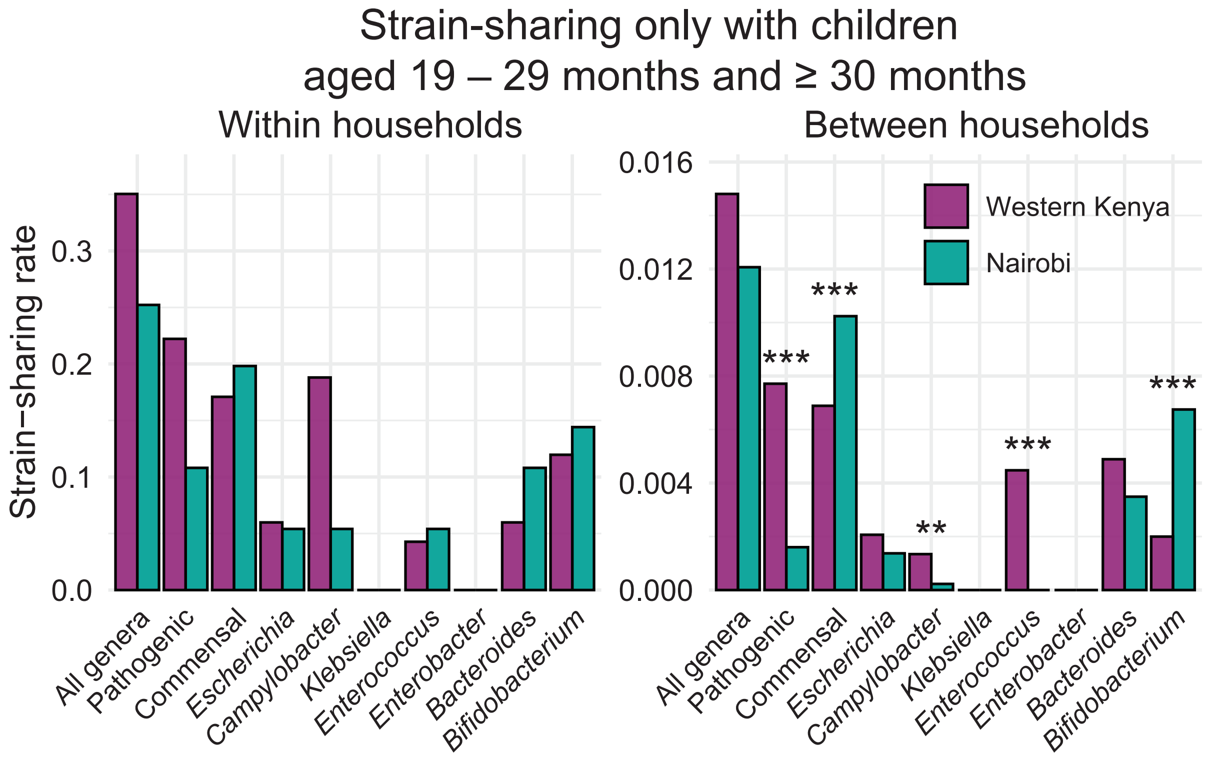


**Fig. S6. Comparison of strain-sharing rates between study sites only with children aged 19 – 29 months and ≥ 30 months.** Statistical significance between groups was assessed using a two-sided non-parametric permutation test (1,000 iterations). (*: $P < 0.05$; **: $P < 0.01$; **: $P < 0.001$).

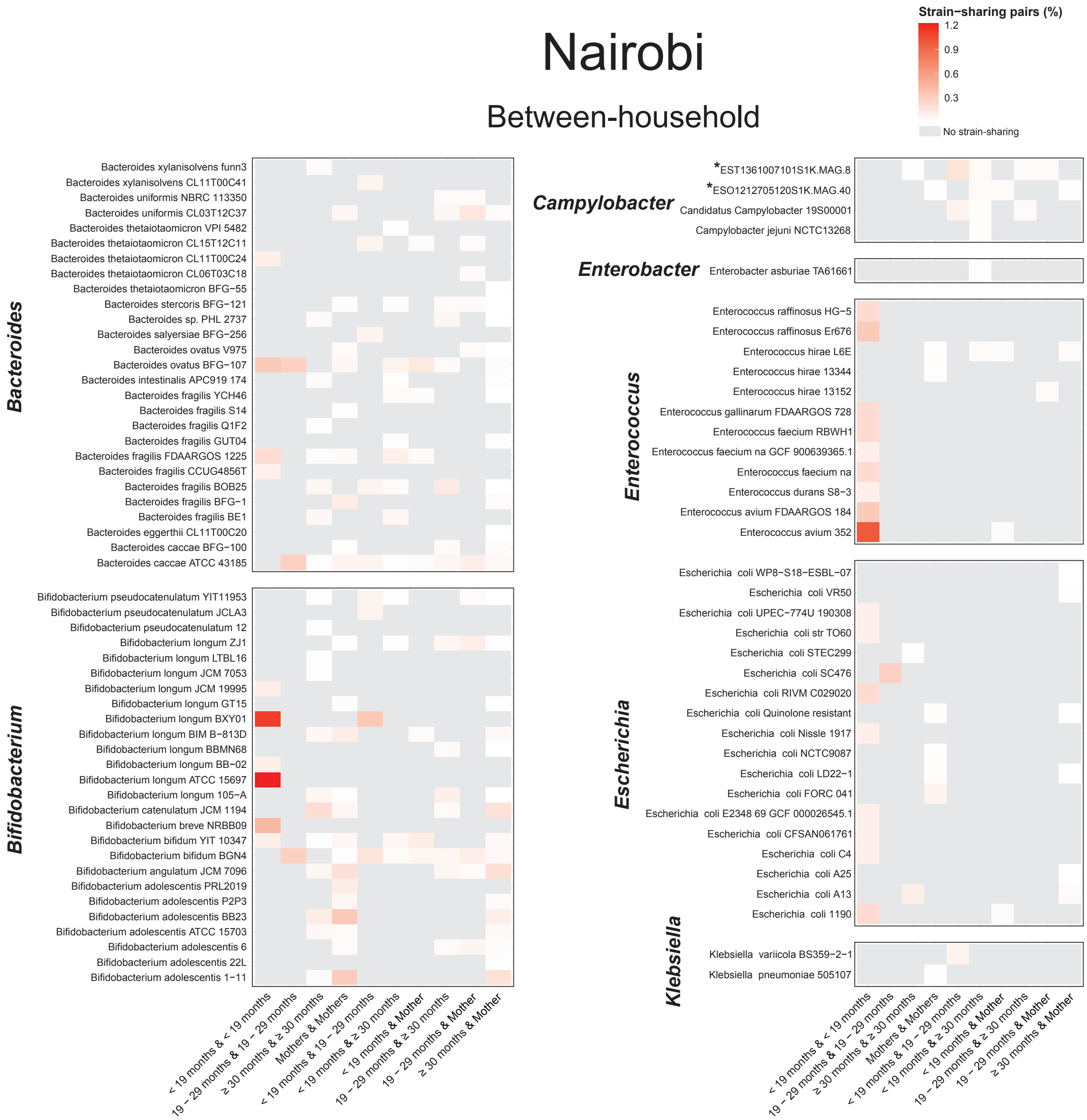


**Fig. S7. Heatmaps of representative reference strains associated with between-household strain sharing in the Nairobi study.** *Campylobacter* strains marked with an asterisk indicate metagenome-assembled genomes reconstructed in this study.

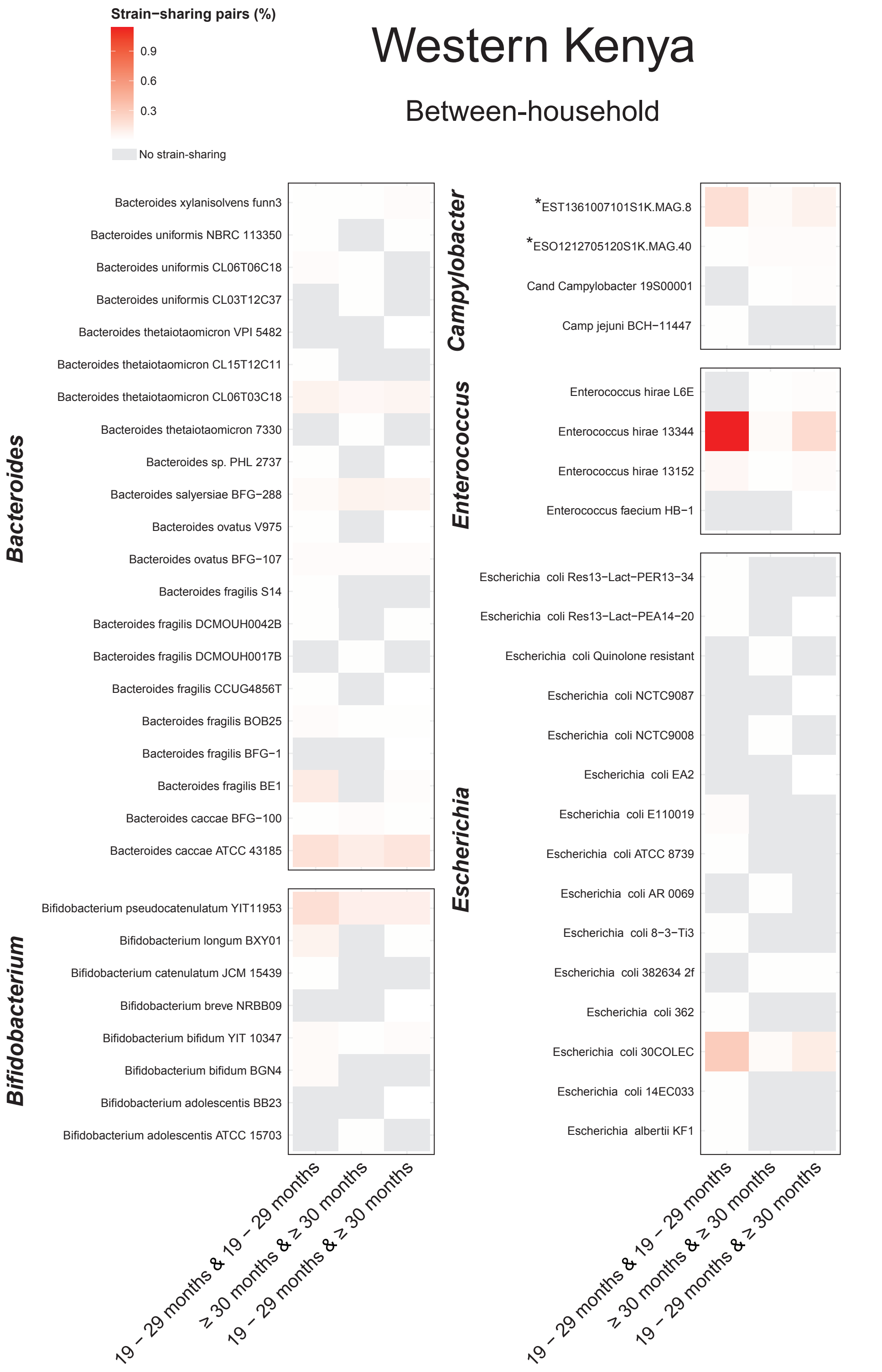


**Fig. S8. Heatmaps of representative reference strains associated with between-household strain-sharing in the Western Kenya study.** *Campylobacter* strains marked with an asterisk indicate metagenome-assembled genomes reconstructed in this study.

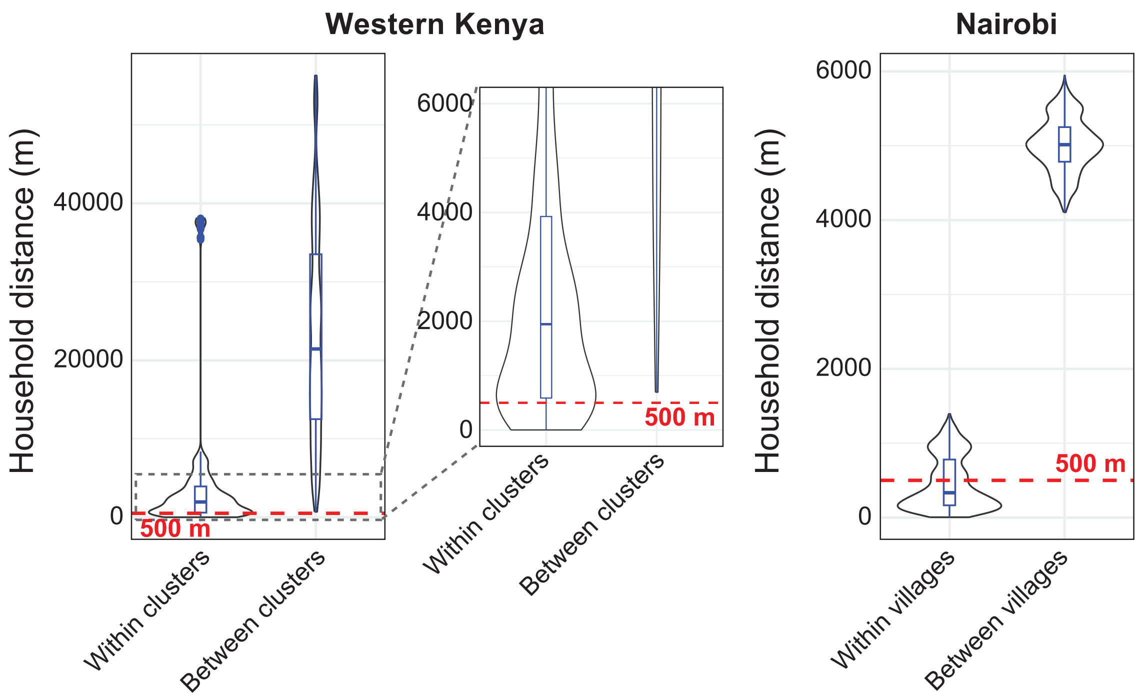


**Fig. S9. Distribution of distances between households in each study site.** Violin plots indicate the density of the distribution, and box plots show the median and interquartile ranges.

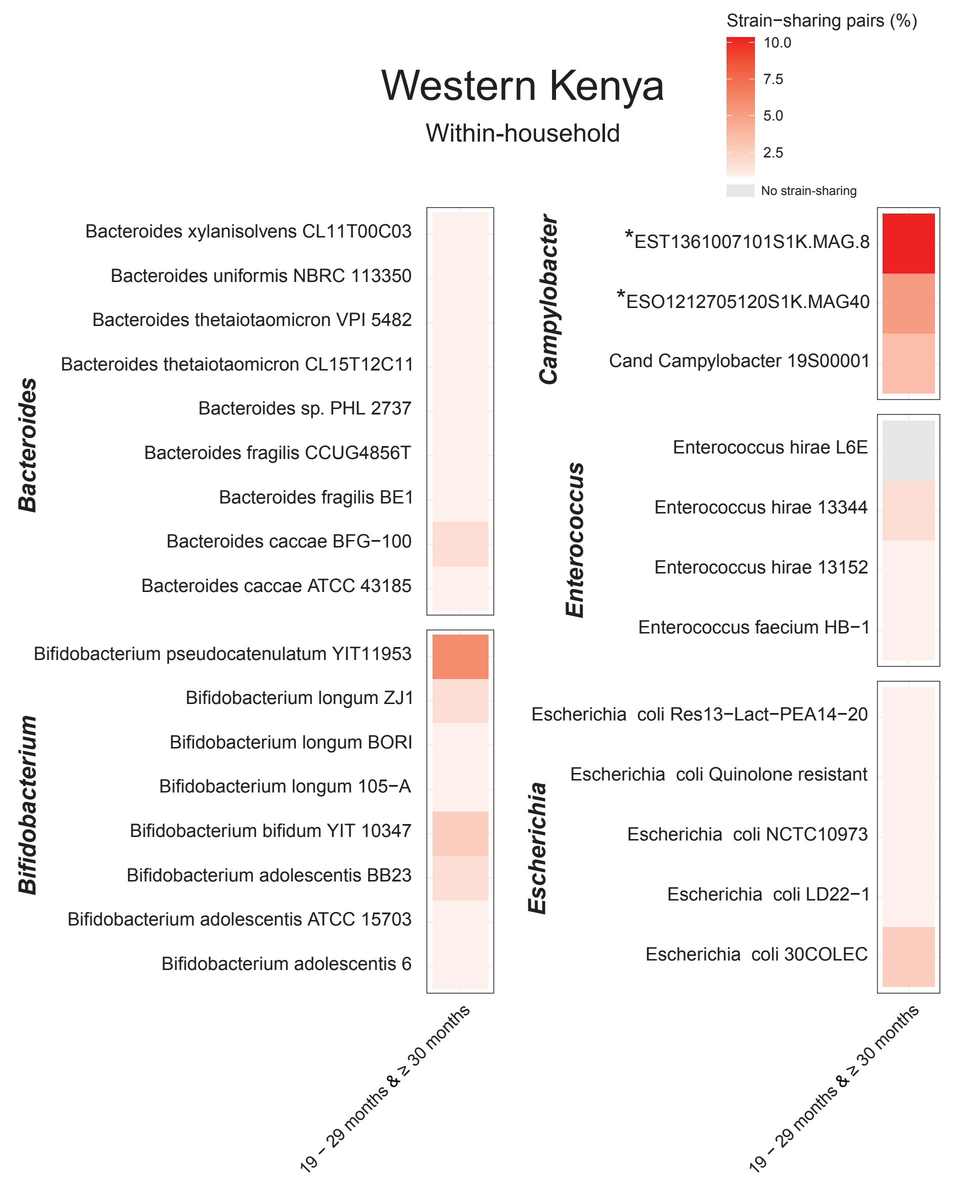


**Fig. S10. Heatmaps of representative reference strains associated with within-household strain-sharing in the Western Kenya study.** *Campylobacter* strains marked with an asterisk indicate metagenome-assembled genomes reconstructed in this study.

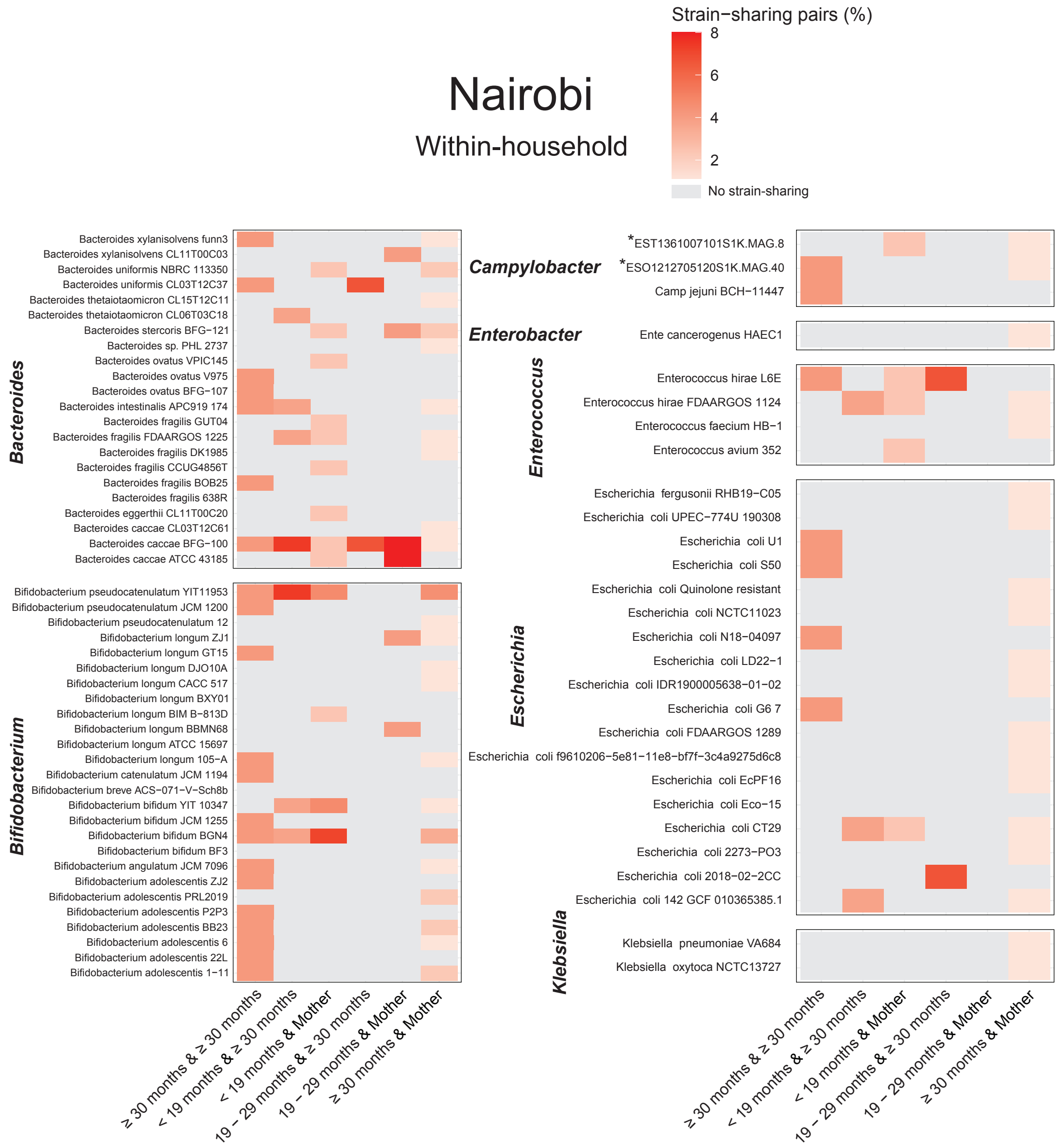


**Fig. S11. Heatmaps of representative reference strains associated with within-household strain-sharing in the Nairobi study.** *Campylobacter* strains marked with an asterisk indicate metagenome-assembled genomes reconstructed in this study.

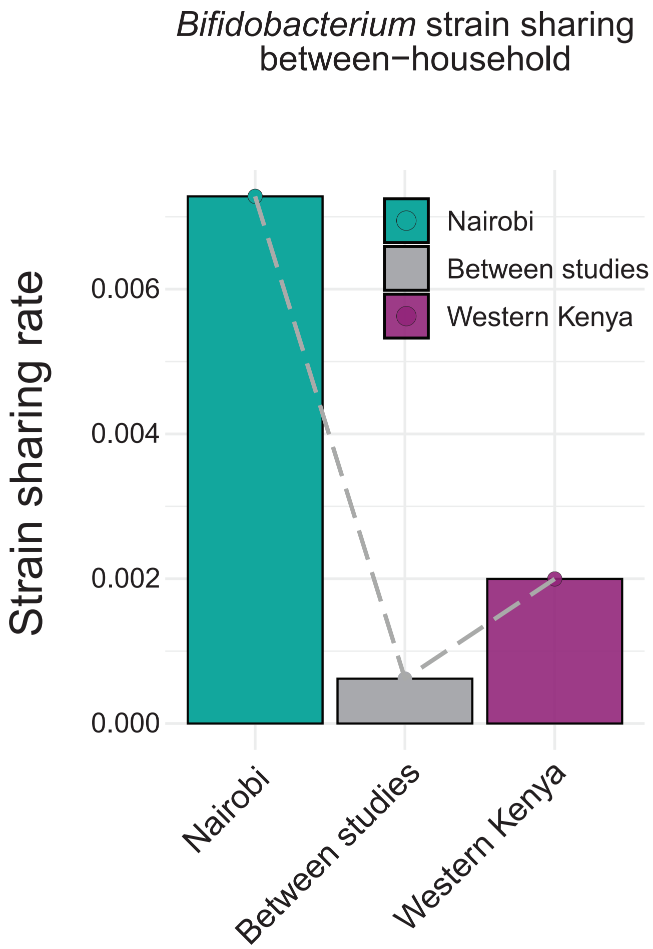


**Fig. S12. Between-household Bifidobacterium strain-sharing within and between studies.** Strain-sharing rate was defined as the average proportion of total sample pairs between households that shared a strain. Statistical significance between groups was assessed using a two-sided non-parametric permutation test (1,000 iterations).

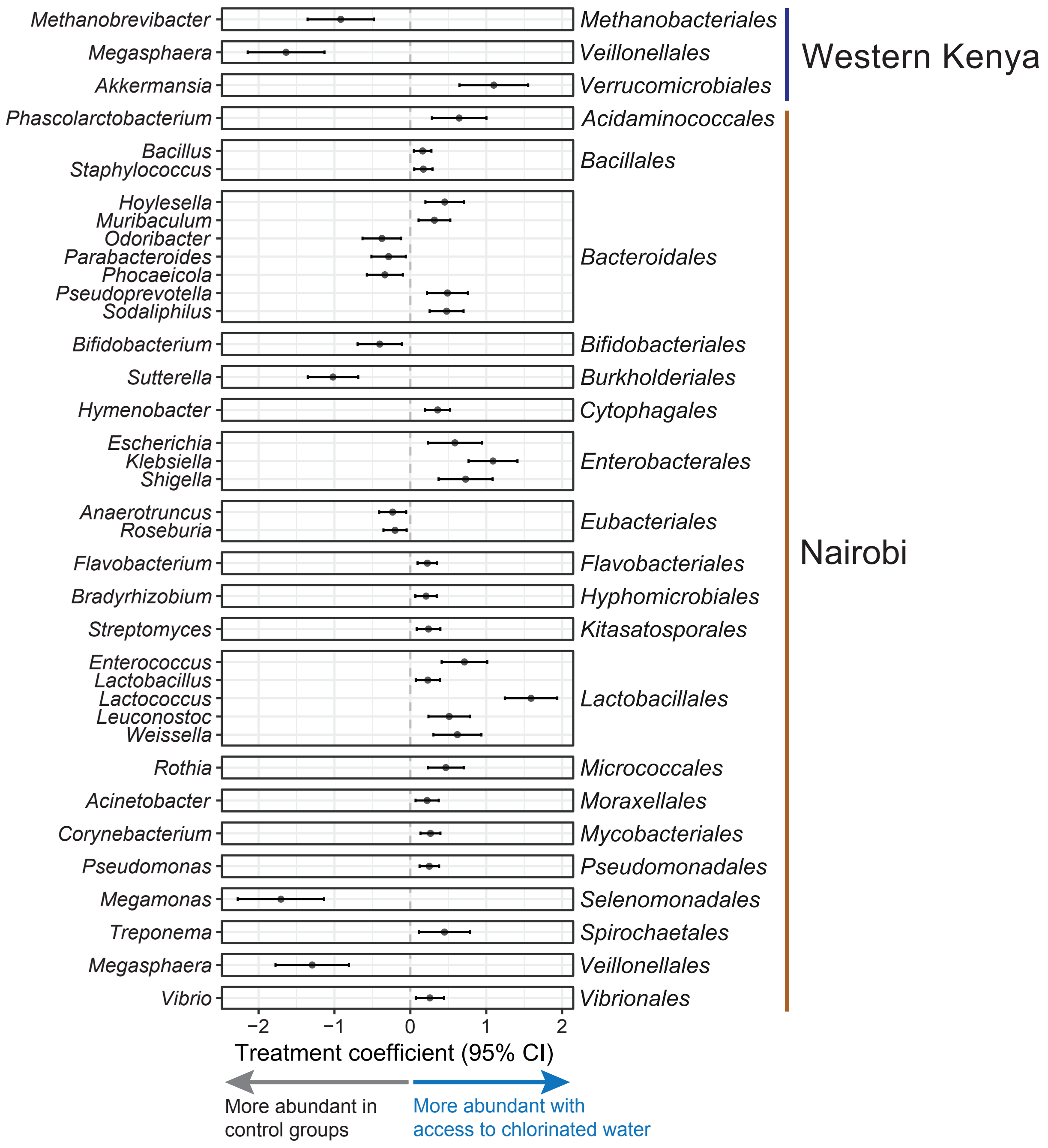


**Fig. S13. Differentially abundant genera by water treatment status, controlling for age group.** Positive coefficients indicate higher abundance in the treatment group, while negative coefficients indicate lower abundance. Error bars represent 95% confidence intervals around the estimated treatment effects.

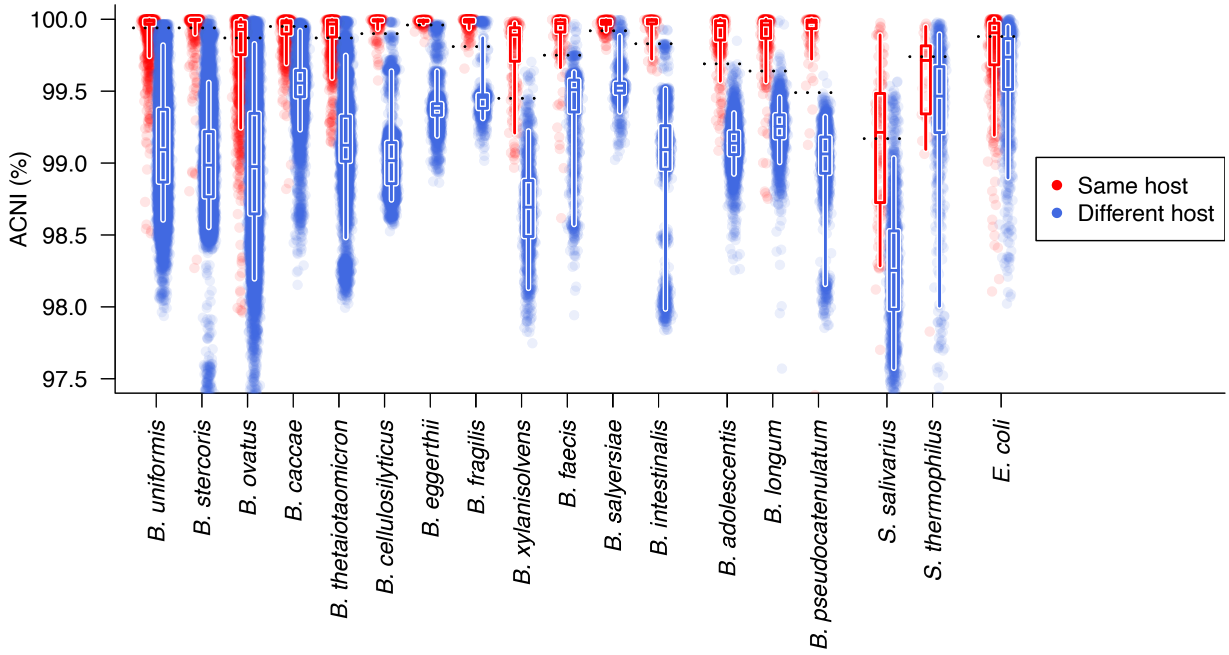


**Fig. S14. Species-specific average callable nucleotide identity (ACNI) thresholds for defining strain-sharing events in the training datasets.** For each species with more than 100 pairwise sample comparisons in the training datasets, the ACNI threshold (dashed horizontal lines) was selected by optimizing classification of samples as same-host (red) or different-host (blue), based on maximization of the F1 score.

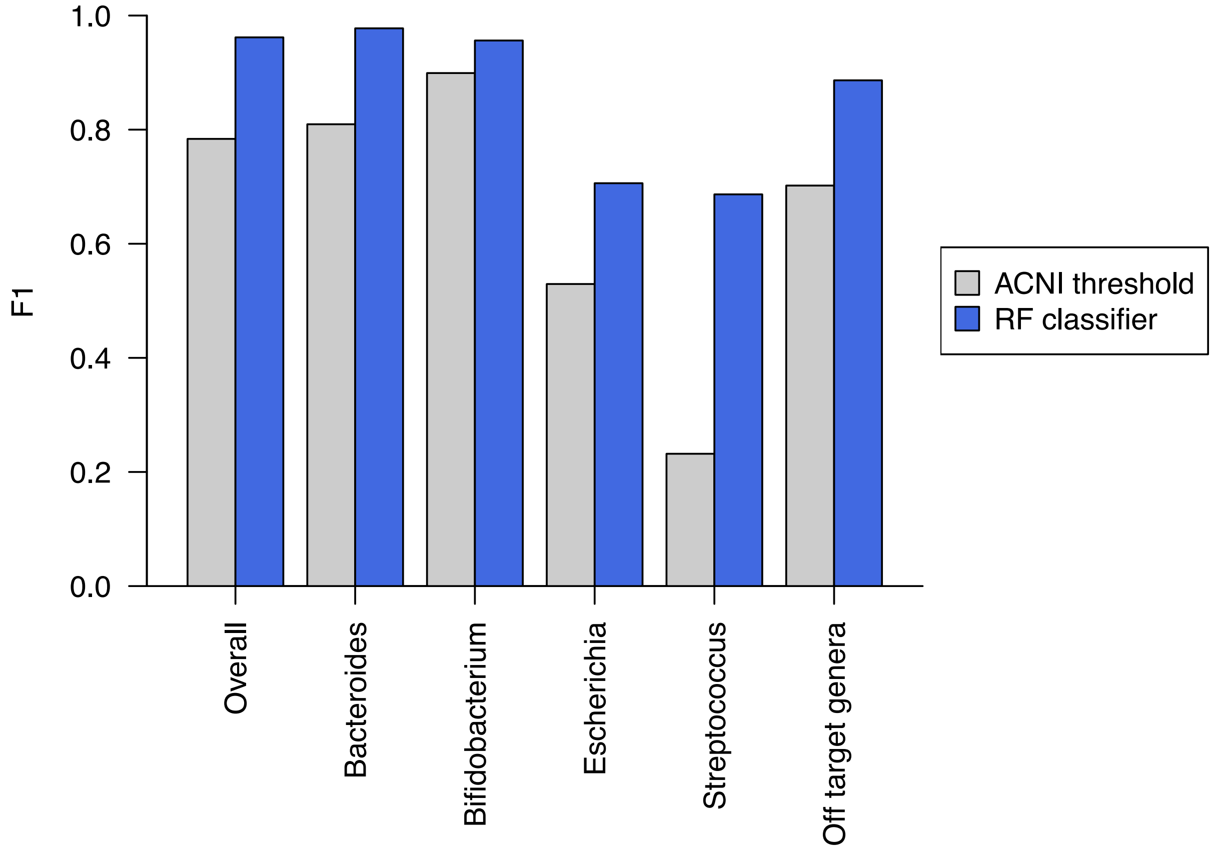


**Fig. S15. Performance of the combined random forest classifier for predicting strain sharing events.** The classifier was trained on 50% of the 135,434 total pairwise strain comparisons and evaluated on the remaining 50% from the traveler and UTI datasets. The bar plot shows F1 scores for all target species included in training ('Overall'), for prevalent genera in the training datasets, and the 13 off-target species not used for training ('Off target genera').

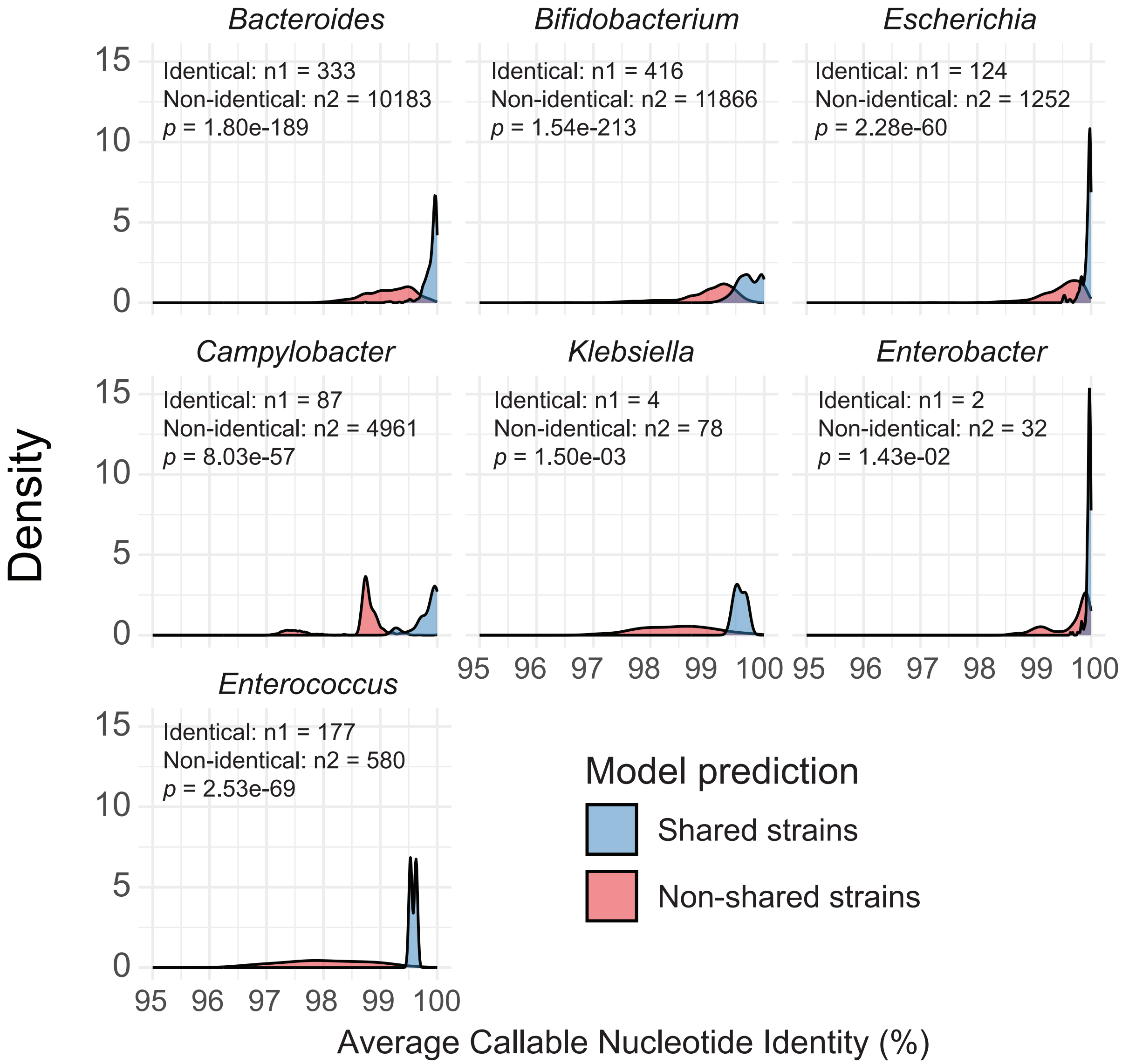


**Fig. S16. Distribution of average callable nucleotide identity (ACNI) values between sample pairs sharing the same StrainGE reference strain, classified as either shared or non-shared strains by a machine learning model.** Each panel corresponds to a different genus. $n_1$ and $n_2$ indicate the number of pairs predicted as identical and non-identical strains. Statistical comparisons were performed using two-sided Mann–Whitney U tests.
